# Hospital Evidence on the Health Impacts of Cumulative Heat

**DOI:** 10.64898/2025.12.11.25342068

**Authors:** Katharina Ledebur, Katharina Brugger, Andrea Schmidt, Marianne Bügelmayer-Blaschek, Martin Schneider, Andrea Hochebner, Peter Klimek

## Abstract

Heatwaves are becoming more frequent, intense, and prolonged. High temperatures raise mortality and morbidity, particularly in older adults and those with pre-existing conditions. However, it remains unclear whether increases in deaths during hot periods are accompanied by similar changes in hospital admissions. Understanding the full health burden of heat requires assessing both outcomes under a unified exposure framework.

We analyzed national hospital data from Austria (2007–2019) using Poisson generalized linear models to estimate the effects of heat on daily admissions and deaths. We defined exposure as the 14-day rolling sum of heat days across various temperature thresholds.

Exposure to 14 days above 30°C increases the relative risk of in-hospital death by 22*·*3% (95% CI 16*·*4-28*·*5%), with larger effects among individuals aged 75+. The affected diagnostic groups during deadly stays include respiratory diseases (129*·*7%, 95% CI 97*·*0–167*·*8), bacterial and vector-borne infectious diseases (41*·*4%, 95% CI 20*·*2–66*·*3), and endocrine, nutritional and metabolic diseases (39*·*7%, 95% CI 8*·*3–80*·*2). The effect for all-cause hospital admissions is negative at -4*·*6% (95% CI -5*·*0 -4*·*01%). However, increases are observed for specific diagnoses, including infectious bacterial and vector-borne infectious diseases (14*·*6%, 95% CI 11*·*3–18*·*1), endocrine, nutritional and metabolic diseases (13*·*3%, 95% CI 9*·*7–17*·*0),and viral, fungal, and parasitic diseases (4*·*6%, 95% CI 1*·*9–7*·*5).

These findings demonstrate that heat waves are associated with higher in-hospital mortality rates across diagnostic groups, with the most vulnerable experiencing the greatest increase in admissions. These results challenge the assumption of uniform increases in hospital demand and underscore the need for targeted action plans.

## 1 Introduction

As the climate warms, heatwaves are becoming more frequent, more intense, and longer-lasting [1]. In Austria temperature extremes (e.g., heat days with daily maximum air temperature > 30°C) have more than doubled in recent decades, for example in Vienna from 9.9 (1961–1990) to 22.0 (1991–2020). Correspondingly, heat-related hospitalizations are about 27% higher in extremely warm summers compared to average summers [2]. Climate projections indicate that mean air temperature will continue to rise, with further increases in the frequency and severity of heat extremes [3].

These trends pose a growing threat to human health. High ambient temperatures and extreme heat events are associated with a variety of negative health outcomes. In 2022, over 60,000 heat-related deaths were estimated across Europe, and in 2023 a further 47,690 deaths were recorded, marking the two highest burdens in recent years [4]. In addition to increased mortality, heat has been linked to higher rates of emergency room visits and hospital admissions, particularly for cardiovascular and respiratory diseases. Mental health crises, pregnancy complications, adverse birth outcomes, and increased medication toxicity have also been reported [5]. Additionally, heat can compromise working conditions and safety for healthcare staff, leading to difficulties in managing heat risks [6].

In Austria, early studies reported significant excess mortality during heat events [7], [8], while more recent analyses suggest partial adaptation, as mortality risks have slightly declined despite a doubling of heatwave frequency and tropical nights [9].

While several studies report increases in hospitalizations during heatwaves, most are restricted to specific conditions or single urban centers [10], [11], [12], [13], [14], [15], [16]. Recent nationwide analyses have documented heterogeneous, diagnosis-specific admission risks across Spain and Portugal [17], [18], while system-wide analyses suggest that cumulative heat effects on inpatient admissions may be offset by reductions in the days following extreme heat [19], [20]. This inconsistency partly reflects differences in exposure metrics, admission types, and whether mortality and morbidity are assessed jointly. No study has jointly examined cause-specific inpatient admissions and in-hospital mortality within a single nationwide hospital dataset, making it difficult to determine whether the divergence between mortality and morbidity risks observed in other settings [21] extends to the inpatient context specifically.

This gap has direct implications for the preparedness of health systems. Overestimating admission surges can result in wasted resources, while underestimating diagnosis-specific vulnerabilities can lead to preventable harm. Furthermore, most heat-health warning systems continue to rely on single-day thresholds, overlooking the cumulative stress resulting from prolonged exposure to heat.

To address this, we conduct a nationwide analysis of hospital records in Austria from 2007 to 2019. We examine how cumulative heat exposure affects daily hospital admissions and in-hospital mortality across diagnostic groups, age strata, sex, and regions. We define heat exposure as the cumulative number of heat days (with maximum air temperature above a given threshold between 26 and 32°C) within the previous 14 days, applying nonlinear Poisson regression models to quantify relative risks. Using a unified exposure definition for both in-hospital mortality and admission, as well as applying consistent stratification, allows us to provide a comprehensive assessment of the differential burden of heat on health outcomes. This approach enables direct comparisons across conditions and populations, yielding insights that are essential for designing effective heat-health adaptation strategies.

## 2 Methods Study region

Austria lies in Central Europe with predominantly temperate climate (Cfb) and alpine/snow climates at higher elevations (Dfb, Dfc, ET, EF) [22]. Analyses are conducted at the 32 care regions (“Versorgungs- regionen”; NUTS-3–comparable). The highest admission and death counts occur in the most populated areas (e.g., Vienna, Graz, Linz, Innsbruck; Figure 1). From 2007 to 2019, hospitals recorded an average of 6,914 admissions and 101 deaths per day during the summer months (Figure 1).

**Figure 1:**
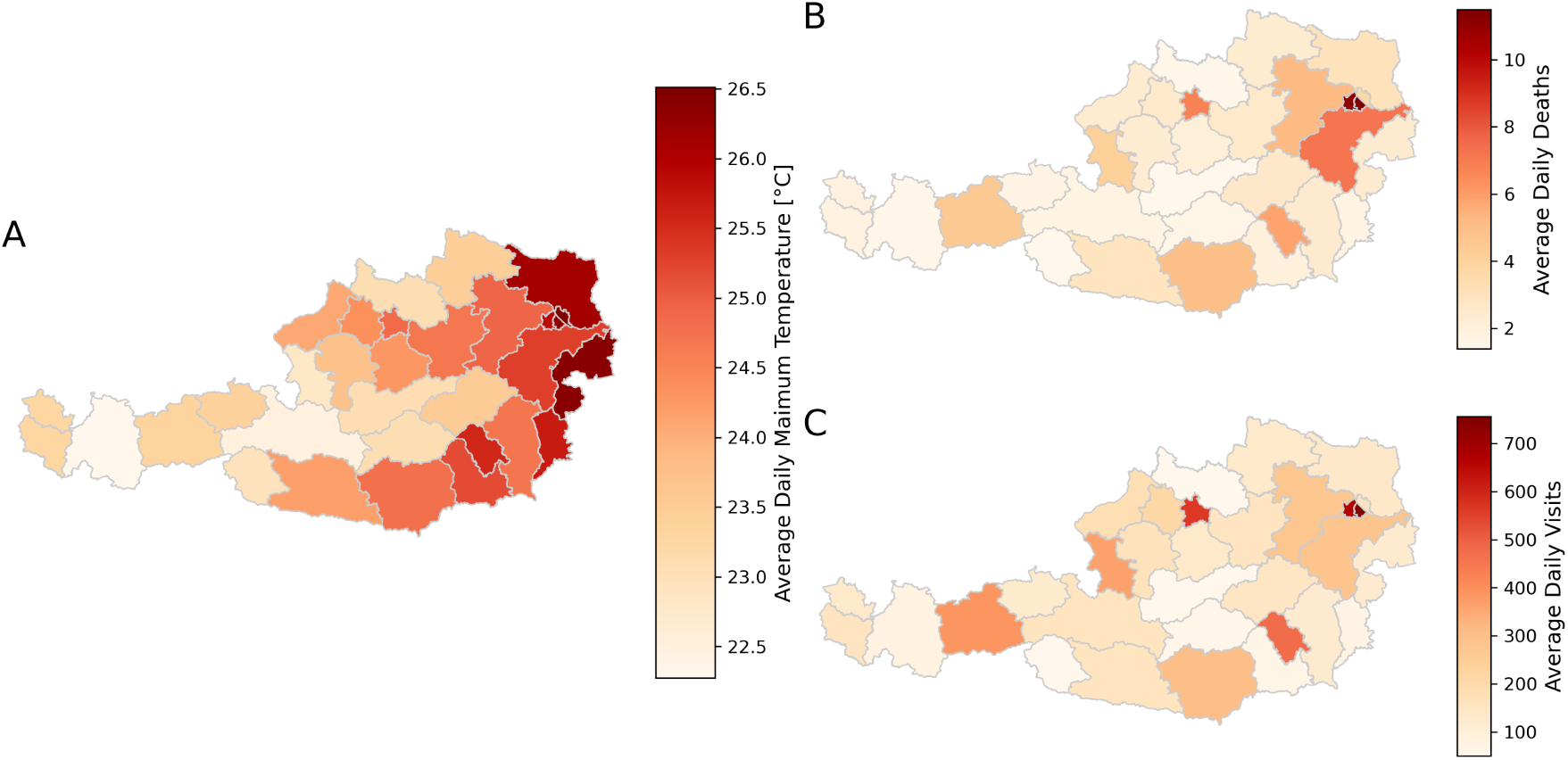
Regional patterns of temperature, hospital admissions, and mortality in Austria. (A) Average daily maximum temperature by region during the study period. (B) Average daily number of in-hospital deaths per region. (C) Average daily number of hospital admissions per region. All values are aggregated at the care-regional level and averaged over the summer months (June, July, and August) over the years 2007-2019, reflecting population and service volume (health system burden).

### Study design and data sources

We performed a retrospective nationwide study for summers 2007–2019. The dataset provided by the Austrian Ministry of Health contains all hospital stays in Austria during the study period, containing one record per stay with admission/discharge dates, age group (5-year bands), sex, region of residence, region of care, primary and secondary ICD-10 diagnoses, and discharge type (including in-hospital death). Main analyses use primary diagnoses; sensitivity analyses include primary and secondary diagnoses in the supplementary information (SI).

Meteorological data were obtained from the largest Austrian private provider of weather forecasts and severe weather warnings (UBIMET). The used reanalysis data provides daily maximum/minimum temperatures on a 1 km^2^ grid; municipality values were population-weighted and aggregated to care regions (population weights).

### Outcomes

We model daily hospital admissions (all stays with that day as admission date) and daily in-hospital deaths (stays discharged as death on that day), overall and by ICD-10 chapters (A–N and O–Z). Models adjust for aggregated age groups (0–44, 45–74, 75+ years), sex, region fixed effects, day of week, and region-specific long-term trends (days since study start). Age groups were aggregated to ensure sufficient statistical power and stable estimates across strata, while retaining meaningful distinctions in heat vulnerability across the life course.

### Exposure and model specification

Heat exposure is the rolling 14-day count of heat days, with thresholds from 26–32 *^◦^*C for daily maximum temperature. The distribution and temporal variation of cumulative heat exposure across summer region-days is shown in SI Figure 6.

To allow for non-linear cumulative effects, the rolling heat exposure *H* was transformed as *H^α^*, where *α* controls the shape of the exposure–response relationship, allowing for diminishing or accelerating effects of repeated heat exposure. The parameter *α* was selected via grid search over a predefined range (0.1 to 3). For each candidate value, a separate model was estimated, and the optimal specification was chosen based on the Akaike Information Criterion (AIC), conditional on statistical significance of the exposure term.

We estimate incidence rate ratios (IRRs) via Poisson regression with robust standard errors, controlling for the covariates above. To capture the effect of consecutive heat days, we also require at least *Z* consecutive heat days within the prior 14 days (tested *Z* = 1–5). Age-, sex-, region-, and diagnosis-specific effects are obtained from separate models. Exposure was assigned based on the patient’s region of residence for hospital admissions (morbidity), and based on the region of care for in-hospital deaths (mortality). Full model specification and selection procedures are provided in the SI.

### Additional Cases (AC)

We translate relative risks into absolute burdens by multiplying baseline daily counts during non-heat periods (defined as zero heat days in the preceding 14 days) by (IRR *−* 1). AC are reported as totals and per 100,000 population (averaged 2015–2019, stratified as in each model).

## 3 Results

### 3.1 Impact of heat days on in-hospital mortality and admissions

#### We find significant associations between cumulative heat exposure and in-hospital mortality and admission

For in-hospital mortality, we find significant associations across all tested temperature thresholds. The strongest increase is observed at *T_max_ >* 32*^◦^*C and a 14-day accumulation window (1*·*446, 95% CI: 1*·*330–1*·*574), corresponding to a 44*·*6% increase in risk (Figure 2A).

**Figure 2:**
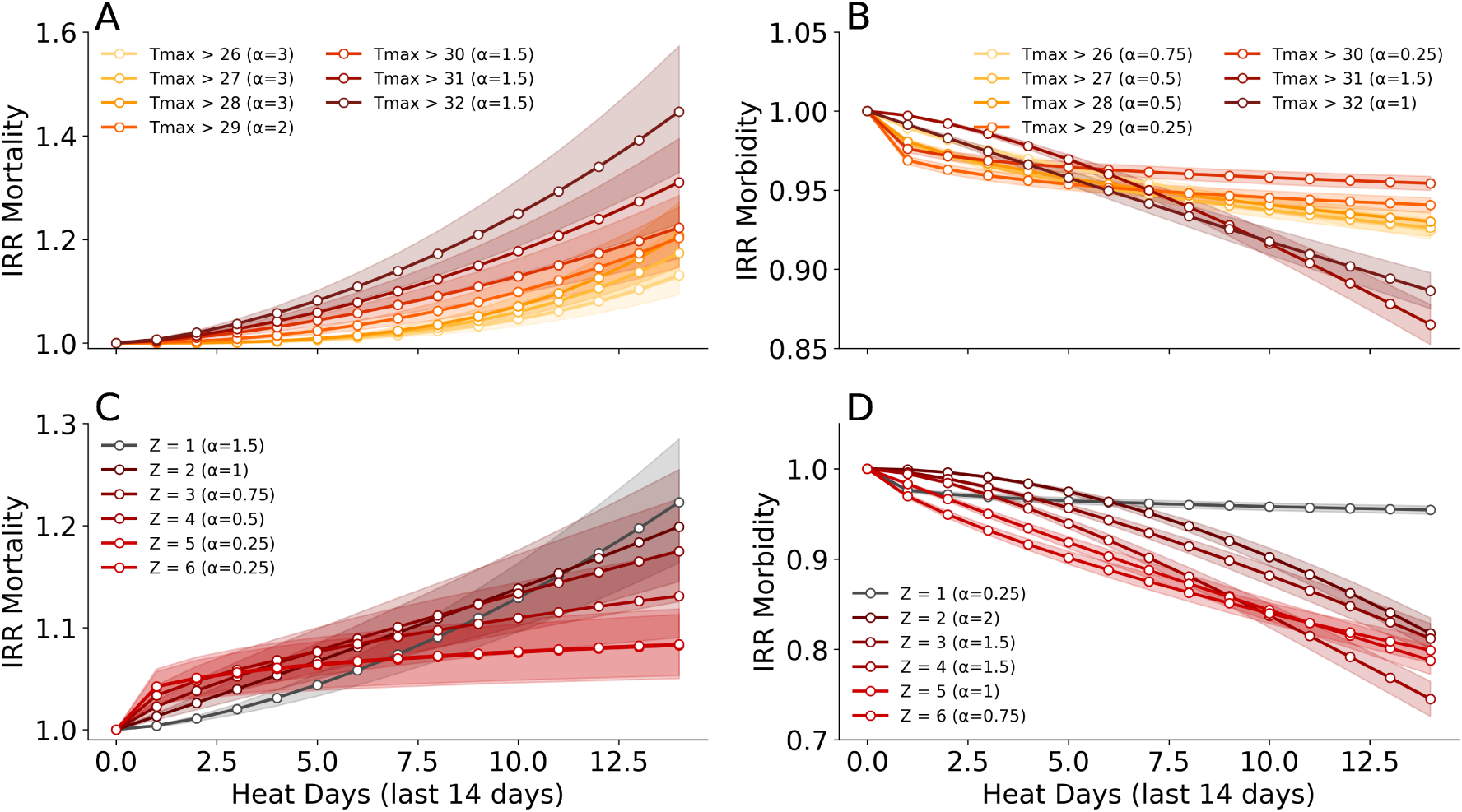
Effects of cumulative and consecutive heat exposure on in-hospital outcomes. Incidence Rate Ratios (IRRs) for in-hospital mortality (A) and admission (B) by threshold of daily maximum temperature (Tmax > 26°C to Tmax > 32°C). (C–D) IRRs by number of consecutive heat days in the past 14 days for in-hospital mortality (C) and admission (D). Shaded bands represent 95% confidence intervals. *α* denotes the scaling parameter optimized for model fit. Note the difference in scales between hospital admissions and in-hospital mortality, respectively. We only show significant results.

For hospital admissions, heat effects are reversed. The highest reduction in relative risk is observed at *T_max_ >* 31*^◦^*C with a 14-day accumulation window (IRR 0*·*865, 95% CI 0*·*852–0*·*878), corresponding to a 13*·*5% decrease in relative risk (Figure 2B).

Model fits indicate that lower thresholds require higher *α* values (steeper nonlinear scaling), suggesting cumulative physiological stress even at milder temperatures. Higher-temperature thresholds require lower *α* values, implying a more linear response at extreme heat. For all-cause admissions, associations remain negative across all tested thresholds in the main 14-day specification.

Consecutive hot days (Z) show similar patterns. In-hospital mortality associations remained elevated under persistence definitions, but effect sizes attenuated as the required number of consecutive heat days increased, while admissions show effects in the opposite direction (Figure 2 C and D, Table 1).

**Table 1:**
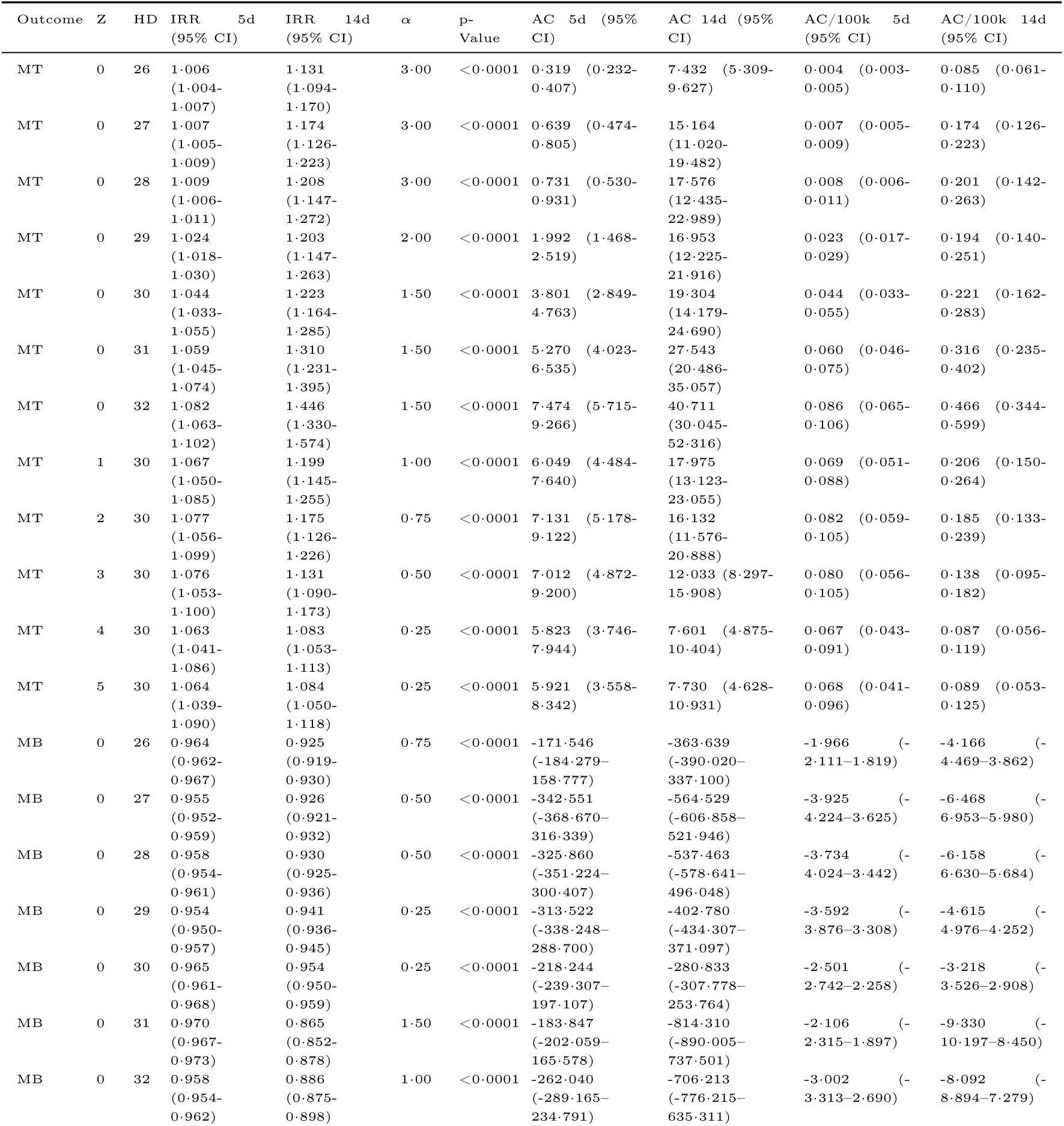
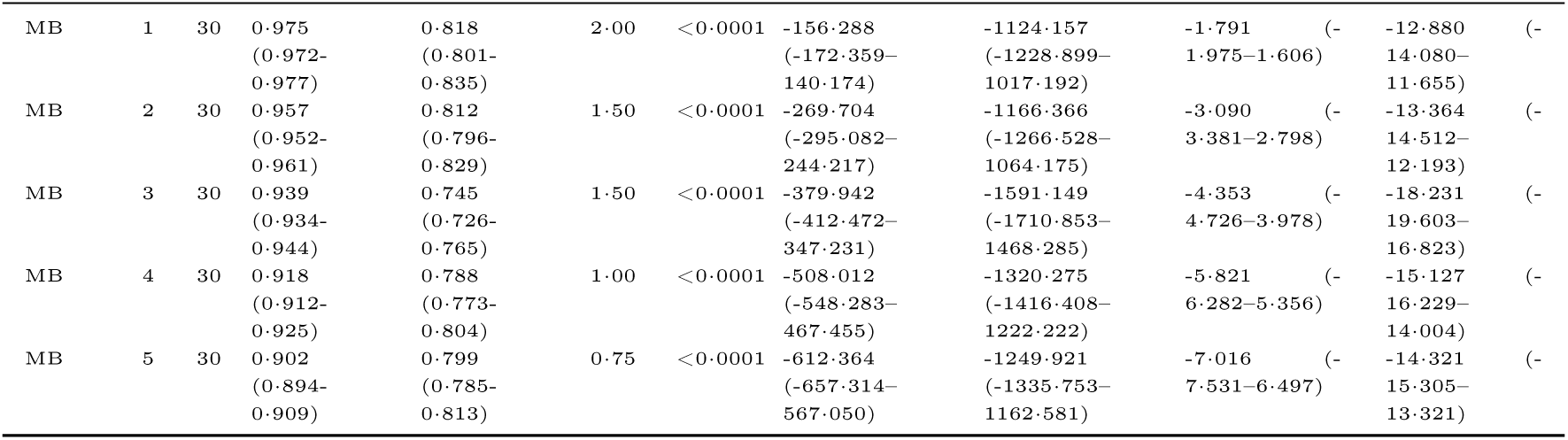
Estimated IRRs and ACs and AC per 100,000 with 95% CI across heat day (HD) thresholds (*T*_max_ *> X^◦^*C) and Z values (consecutive days) for outcome in-hospital mortality (MT) and hospital admissions (MB).

Table 1 summarizes the estimated IRRs, model parameters, and the number of additional in-hospital deaths and admissions per 100,000 people per day.

*IRR*_5*d*_ and *IRR*_14*d*_ denote model-based relative risks evaluated at *H* = 5 and *H* = 14, where *H* is the number of heat days within the preceding 14-day window. That is, they represent hypothetical exposure scenarios with five or fourteen heat days in the past 14 days, rather than averages over observed days. The largest increases in in-hospital mortality risk occur at *T_max_ >* 32*^◦^*C with *IRR*_5*d*_ = 1*·*082 for five heat days and *IRR*_14*d*_ = 1*·*446. These correspond to roughly 7 (or 41) additional in-hospital deaths and 0.09 (or 0.5) additional in-hospital deaths per 100,000 individuals per day. These estimates refer to models without an additional persistence requirement (*Z* = 0). For hospital admissions, the lowest IRRs occur at *T_max_ >* 31*^◦^*C with *IRR*_5*d*_ = 0*·*970 for five heat days and *IRR*_14*d*_ = 0*·*865. These correspond to roughly -184 (or -814) fewer hospital admissions and -2 (or -9) hospital admissions per 100,000 individuals per day.

Stratified analyses show the strongest mortality effects in adults aged 75 years and older (IRR 1*·*294, 95% CI 1*·*217–1*·*377 after 14 heat days), in females compared with males (1*·*360 versus 1*·*178), and regionally in Tyrol, Burgenland, Vienna, and Lower Austria. For admissions, all demographic and regional strata shown in the main stratified analysis have IRRs below 1, with the largest reductions observed among adults aged 75 years and older, males, and in Upper Austria, Tyrol, Salzburg, and Vienna (Figure 13; Table 3, SI).

### 3.2 Diagnosis-specific effects on in-hospital mortality and hospital admissions

#### The effect of heat exposure on in-hospital mortality varies markedly across diagnostic groups (**Figure 3**)

We estimate the association between cumulative heat exposure and in-hospital mortality separately for each ICD-10 chapter (Tmax > 30°C; past 14 days). Across all ages, mortality increases for multiple diagnostic groups. Among these, respiratory diseases show the most pronounced associations (J00–J99: IRR 2*·*297, 95% CI 1*·*970–2*·*678), while increases are also observed for infectious diseases (A00–A99: IRR 1*·*414, 95% CI 1*·*202–1*·*663), endocrine, nutritional and metabolic diseases (E00–E90: IRR 1*·*397, 95% CI 1*·*083–1*·*802), and diseases of the circulatory system (I00–I99: IRR 1*·*272, 95% CI 1*·*068–1*·*515) (Figure 3A).

**Figure 3:**
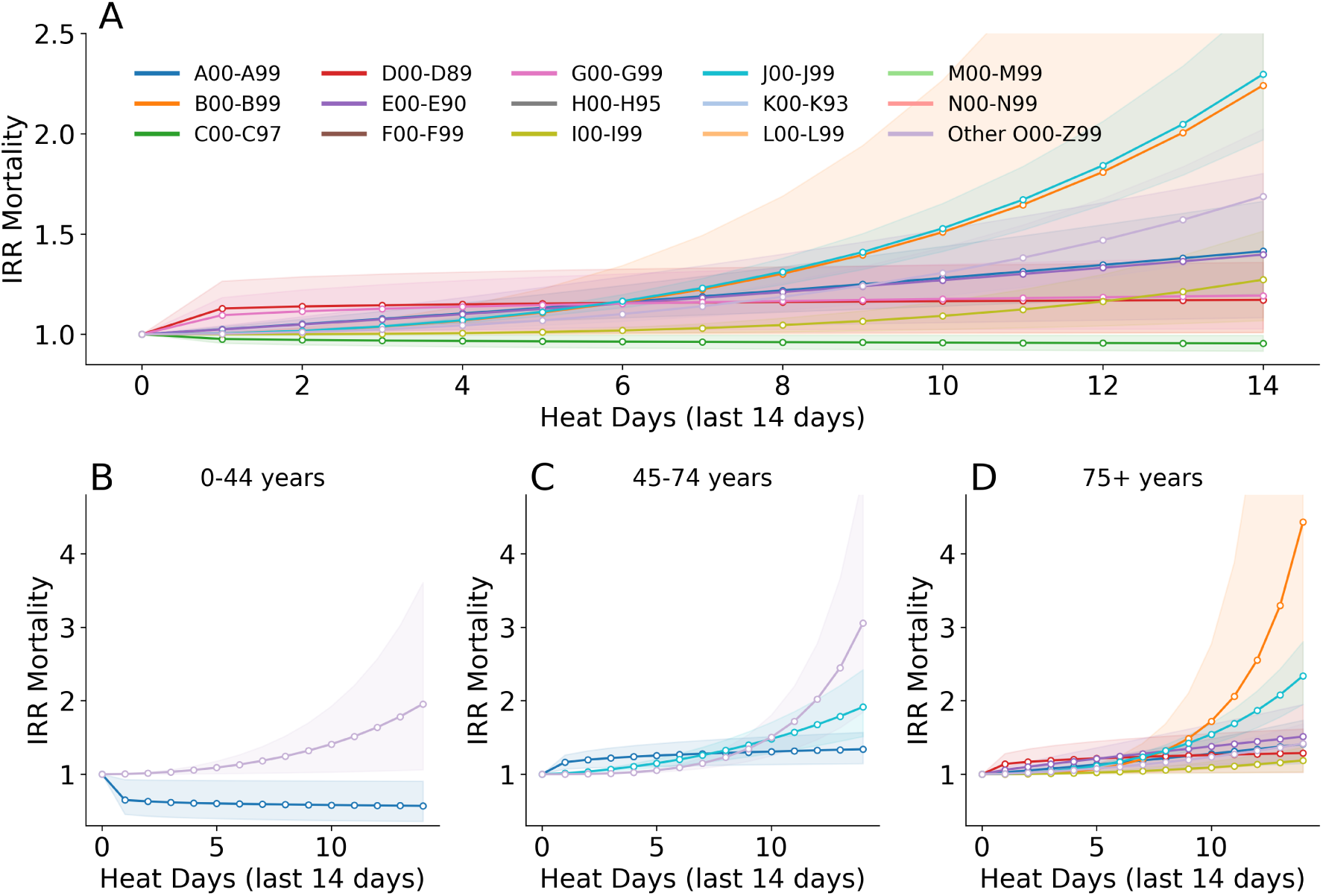
Impact of heat days (Tmax > 30°C) on in-hospital mortality, by diagnosis group and demographic strata. (A) IRRs for in-hospital mortality per diagnosis group (ICD-10 chapters A to N and O–Z combined) associated with the number of heat days (Tmax > 30°C) in the past 14 days. (B–D) IRRs for in-hospital mortality per diagnosis group stratified by age group: 0-44 years (B), 45–74 years (C), and 75+ years (D). Only statistically significant results are shown. Shaded bands represent 95% confidence intervals. Confidence intervals extending beyond the plot range are truncated at the axis limit; full ranges are reported in Table 4 in the SI.

Among age groups, notable differences emerge. In individuals aged 0–44 years, significant associations are observed for certain infectious and parasitic diseases (A00–A99) and diagnoses in the remaining ICD categories (O00–Z99). Among those aged 45–74 years, increased mortality risks are observed particularly for infectious diseases and respiratory conditions. For individuals aged 75 years and older, heat-related mortality increases are more widespread, with elevated risks observed for infectious diseases, endocrine–metabolic diseases, circulatory diseases, and respiratory diseases, the latter showing the largest effect sizes (Figure 3B, C).

Full IRRs, confidence intervals, *α* estimates, and daily additional deaths per 100,000 are reported in Table 4. More granular diagnosis-level patterns are shown in Figure 14 and Table 6 in the SI.

#### Hospital admissions also show diagnosis-specific variation in response to cumulative heat exposure, though the relative increases are generally smaller than those observed for inhospital mortality (**Figure 4**)

Across all ages combined, the largest increases occur for certain infectious and parasitic diseases (A00–A99, B00–B99) and endocrine, nutritional and metabolic diseases (E00–E90), while many other diagnostic groups show decreases. The largest relative increases are observed for stays involving endocrine, nutritional and metabolic diseases (E00–E90: IRR 1*·*133, 95% CI 1*·*097–1*·*170) (Figure 4A).

**Figure 4:**
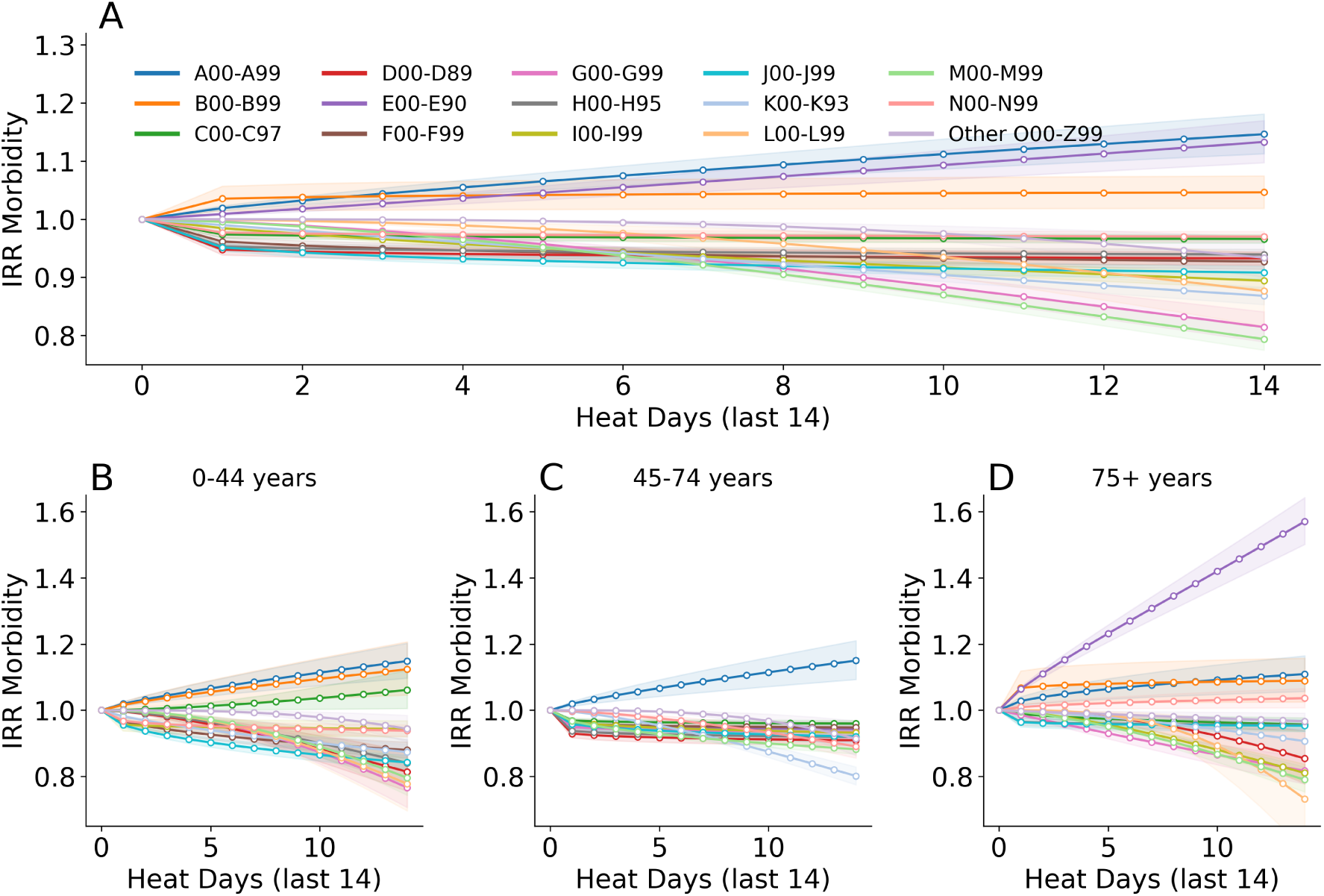
Impact of heat days (Tmax > 30°C) on hospital admissions, by diagnosis group and demographic strata.(A) IRRs for hospital admissions per diagnosis group (ICD-10 chapters A to N and O–Z combined) associated with the number of heat days (daily maximum temperature above 30 degrees) in the past 14 days. (B–D) IRRs for hospital admissions per diagnosis group associated with the number of heat days stratified by age group: 0-44 years (B), 45–74 years (C), and 75+ years (D). Only statistically significant results are shown. Shaded areas represent 95% confidence intervals. Confidence intervals extending beyond the plot range are truncated at the axis limit.

The age pattern mirrors that observed for mortality. Among individuals aged 0–44 years, increases are observed for certain infectious and parasitic diseases (A00–A99, B00–B99) and neoplasms (C00–C97). In adults aged 45–74 years, increases are primarily observed for infectious diseases (A00–A99). In the 75+ age group, increases are most pronounced for endocrine, nutritional and metabolic diseases (E00–E90), with smaller increases also observed for certain infectious and parasitic diseases (A00–A99, B00–B99) and diseases of the genitourinary system (N00–N99) (Figure 4C, D).

Full IRRs, confidence intervals, *α* parameters, and estimated additional daily admissions per 100,000 population are provided in Table 5. More detailed diagnosis-level patterns are shown in Figure 15 and Table 7 in the SI.

#### Reductions in hospital admissions and in-hospital mortality

For in-hospital mortality, we observe small but statistically significant decreases for neoplasms (C00–C97), with IRRs below 1 across all ages, and for circulatory diseases (I00–I99) and skin diseases (L00–L99) in older adults.

Declines in hospital admissions are more widespread and involve multiple diagnostic groups, including neoplasms (C00–C97), diseases of the blood and immune system (D00–D89), mental and behavioral disorders (F00–F99), diseases of the nervous system (G00–G99), circulatory diseases (I00–I99), respiratory diseases (J00–J99), digestive diseases (K00–K93), and musculoskeletal diseases (M00–M99).

#### Comparing the effects of diagnoses across relative and absolute metrics reveals differences in the nature of heat-related health impacts (**Figure 5**)

Relative risks are generally higher for in-hospital mortality than for hospital admissions, suggesting that heat impacts clinical severity more than hospital admission.

**Figure 5:**
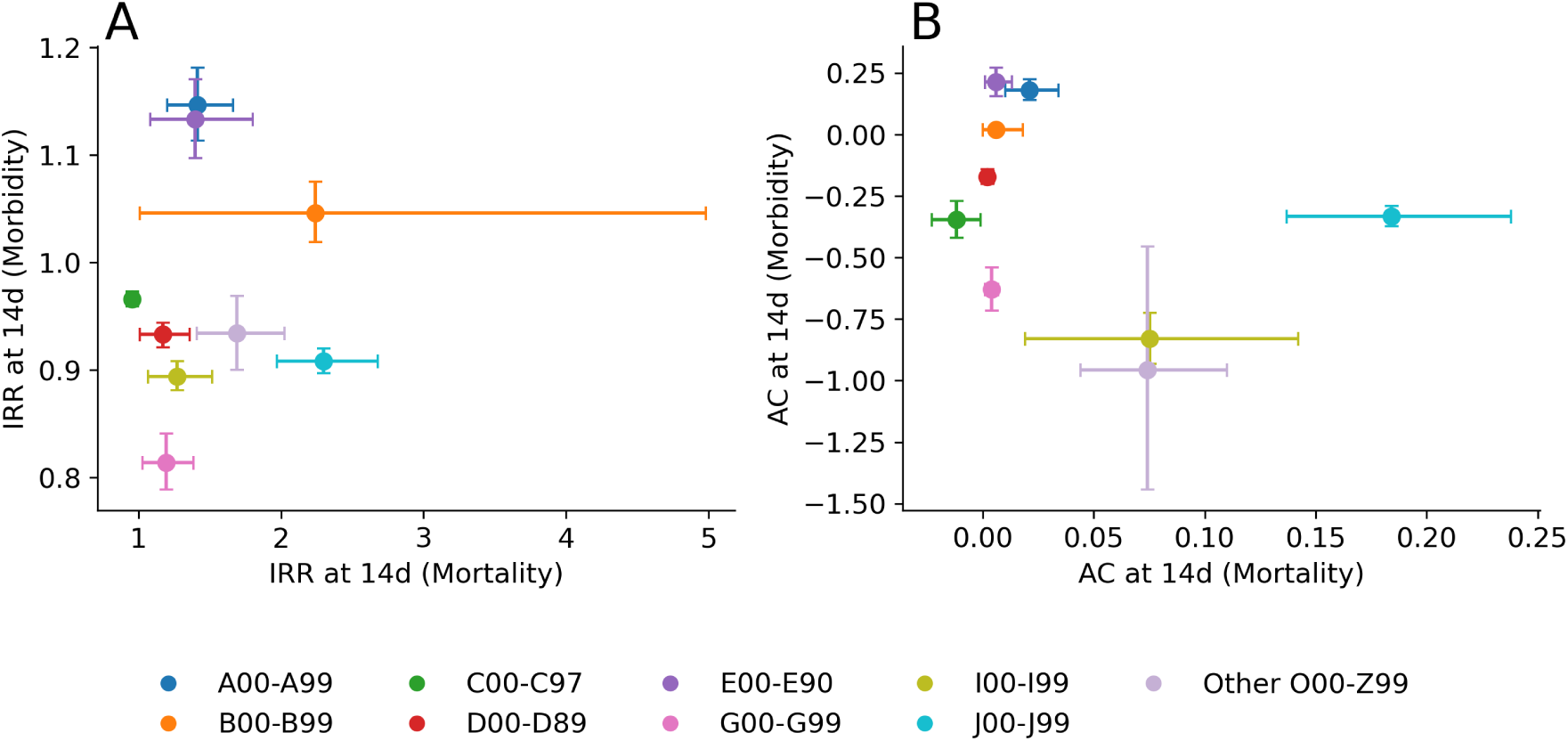
Comparative effects of 14 consecutive heat days (Tmax > 30°C) on in-hospital deaths and hospital admissions across ICD-10 diagnosis groups. (A) IRRs for in-hospital deaths vs. hospital admissions and (B) Additional Cases (AC) per 100,000 per day for in-hospital deaths vs. hospital admissions. Each point represents a diagnosis group (color-coded by ICD-10 chapter). We only show significant effects for both outcomes. Error bars indicate 95% CI.

There were significant increases in in-hospital deaths and admissions for endocrine and metabolic diseases, infectious and parasitic diseases. Respiratory diseases, diseases classified in the O00–Z99 range, circulatory, and nervous system disorders show elevated mortality but no increase in admissions.

The largest excess burdens per 100,000 individuals arise from endocrine and metabolic conditions for hospital admissions and respiratory conditions for in-hospital death.

This suggests that heat does not uniformly increase hospital utilization and deaths, but rather shifts the clinical case mix and the conditions under which patients arrive and deteriorate within the hospital system.

### 3.3 Sensitivity analyses

Sensitivity analyses supported the main findings. Results were directionally consistent when using 7-day and 21-day accumulation windows (SI Figure 7, Figure 8). Heat definitions combining maximum temperature and relative humidity produced similar patterns to temperature-only thresholds (SI Figure 9).

Repeating the diagnosis-specific analysis with both primary and secondary diagnoses preserved the patterns reported above. Effect sizes were larger under the broader case definition, reflecting the inclusion of comorbid conditions documented during the stay (SI Figure 16, Figure 17, Table 8, Table 9).

Our primary analysis uses a rolling accumulation metric, which imposes uniform weighting across a fixed window but allows direct interpretation in terms of heat days and translates into additional cases under interpretable scenarios such as five or fourteen heat days. Distributed lag non-linear models (DLNMs) were fitted as an additional check of the lag structure [4], [23], [24], [25] (see SI). For in-hospital mortality, the cumulative exposure-response increases monotonically above approximately 22°C, with a cumulative IRR of 1*·*101 (95% CI 1*·*077–1*·*127) at 30°C relative to 20°C (Figure 11). The lag-response is concentrated at lag 0–2 and approaches null beyond lag 5, supporting the rolling-window assumption that recent days carry most of the signal. For hospital admissions, the DLNM shows a modest positive association at extreme temperatures (IRR 1*·*019, 95% CI 1*·*015–1*·*024 at 30°C versus 20°C)(Figure 12), in contrast to the negative association observed for sustained cumulative heat exposure in the primary model. This difference is expected because the DLNM captures acute effects of daily temperature, whereas the primary model captures the net association of prolonged heat periods, including possible changes in elective care, admission practices, and care-seeking.

### Timing of in-hospital death

Heat-related risk was higher for deaths occurring on the day of admission than for deaths occurring later in the stay (Figure 10). This pattern suggests that acute deterioration or delayed presentation may contribute to the in-hospital mortality signal during sustained heat periods.

## 4 Discussion

In this nationwide study of Austrian hospital records from 2007 to 2019, cumulative heat exposure is associated with higher in-hospital mortality but not with a uniform increase in hospital admissions. In-hospital mortality increases sharply during heat periods, with a 29*·*4% higher risk among adults aged 75 years and older after 14 heat days above 30°C. The strongest diagnosis-specific mortality associations were observed for respiratory diseases, infectious and parasitic diseases, and endocrine, nutritional, and metabolic diseases. In contrast, all-cause admissions declined during sustained heat periods, although admissions increased for selected diagnoses, particularly infectious diseases, endocrine and metabolic diseases, and genitourinary diseases in older adults.

This divergence between deaths and hospital admissions is the central finding of our study. It suggests that heat does not simply increase hospital demand across the board. Rather, sustained heat appears to change the composition and severity of patients reaching hospital care. Some conditions show increased admission burden, whereas others show reduced inpatient activity, potentially reflecting postponed elective care, altered care-seeking, changes in admission thresholds, or shifts in diagnostic coding during heat periods. At the same time, the consistent increase in in-hospital mortality indicates that patients who are admitted during heat periods may be more clinically vulnerable or may deteriorate more rapidly.

### Cause-specific in-hospital mortality

The largest mortality increases were observed for respiratory diseases, infectious and parasitic diseases, and other diagnoses (O00-Z99). For example, mortality among patients with endocrine and metabolic diseases increased by more than 40% under prolonged heat exposure. Respiratory diseases showed particularly strong associations, with mortality more than doubling after 14 days of sustained heat exposure. Infectious and parasitic diseases also showed elevated risks, although the estimates for some subgroups were accompanied by wider confidence intervals.

Cause-specific heat mortality has been reported across a broad set of diagnoses, but most studies targeted selected causes rather than a comprehensive cause-wide screen. Nonetheless, our findings align with broader patterns in the literature. Studies have consistently reported excess heat-related deaths for cardiovascular and respiratory causes, with additional signals for endocrine or metabolic, infectious, mental, and nervous system disorders in settings that examined a broader set of diagnoses [18], [26], [27], [28], [29], [30], [31].

### Cause-specific hospital admissions

Heat-related increases in hospital admissions were smaller and more heterogeneous. Increases were primarily observed for endocrine, nutritional and metabolic diseases, infectious and parasitic diseases, and, in older adults, genitourinary diseases. For example, admissions for endocrine and metabolic diseases rose by 13% after 14 heat days, with even larger increases among adults aged 75 years and older.

Our findings are partly consistent with earlier work showing that heat-related morbidity is most evident for certain diagnoses, though many studies have focused on specific causes. Compared with these studies, our diagnosis-specific estimates are generally smaller in magnitude and more heterogeneous. In the United States, extreme apparent temperatures were associated with modest increases in total admissions, primarily driven by renal and respiratory diseases [16]. In Adelaide, increases were also concentrated in renal and mental health conditions [15] and in northern Italy, prolonged heatwaves particularly raised hospitalizations for heat-related and respiratory diseases [32]. The 2006 California heatwave led to sharp rises in emergency visits and admissions for heat illness, renal failure, and electrolyte imbalance [11], while more recent work in Glasgow reported higher trauma and fracture presentations during heat and humidity heatwaves [14]. A recent nationwide analysis found an overall rise of nearly 20% in admissions during heatwaves [18]. Achebak *et al.* documented heterogeneous, diagnosis-specific admission risks across 48 Spanish provinces, with the strongest effects for metabolic, renal, and infectious diseases and little increase for cardiovascular admissions [17]. Gould *et al.* similarly found that ED visits and hospitalizations respond differently to heat than mortality, with morbidity effects more broadly distributed across diagnoses and age groups [21].

### Interpreting divergent in-hospital mortality and admission patterns

The observed decreases in relative risks, for instance in neoplasms, are unlikely to reflect a protective effect of heat. They may be due to changes in hospital care and diagnostic coding during periods of heat stress. Postponed procedures and early discharges could shift the case mix toward more severe patients. Differences in care-seeking behavior during hot periods could further reinforce reductions in relative risk. On the other hand, primary diagnoses may shift toward acute, heat-related conditions, such as dehydration or renal failure, which tend to increase during hot periods in Austria [2].

Since mortality outcomes are limited to in-hospital deaths, the displacement of fatal events to out-of-hospital settings could also contribute to the observed decreases. While we do not have data on care-seeking in out-of-hospital settings, our sensitivity analysis points to the possibility that patients presenting late to hospital (i.e. dying on the day of admission) have a particularly high risk of dying in heat periods, compared to other patients, and compared to patients dying in the hospital after a few days.

Ultimately, these patterns are more likely to reflect changes in care-seeking, admission practices, case mix, or coding than true reductions in disease risk.

The distinction between admissions and in-hospital deaths is also important for interpretation. Admissions were linked to heat exposure on the day of hospital entry, whereas deaths were linked to exposure on the day of death. Admission models therefore capture entry into the hospital system, while mortality models capture fatal outcomes among admitted patients. Mortality may rise even when admissions remain stable or decline if admitted patients are more vulnerable, present later, or deteriorate more rapidly. Because our data do not include out-of-hospital deaths, ambulatory care, emergency department visits without admission, or long-term care contacts, we cannot determine how much of the heat-related burden occurs before hospital admission.

This interpretation is consistent with previous evidence showing stronger and more consistent heat effects for mortality than for hospital admissions. Studies from England and Spain found that mortality increased during hot periods while admissions rose only modestly or selectively, a pattern interpreted as consistent with some heat-related deaths occurring before hospital contact [13], [17]. Multicity inpatient evidence also suggests that patients are more likely to die in hospital on hotter days, while admission signals are more diagnosis-specific [33]. Among patients with end-stage renal disease, hospitalizations and deaths increased during periods of extreme heat, though the effect size was smaller for hospital admissions [34]. Recent evidence from California and England shows that morbidity and mortality respond to heat through distinct mechanisms, with mortality concentrated among older adults and specific diagnoses while morbidity effects are broader and more diffuse [20], [21]. Overall, the literature indicates stronger and more consistent effects on mortality than on hospital admissions. Our findings are consistent with this pattern.

### Strengths and limitations

A key strength of this study is the use of a nationwide hospital dataset covering more than nine million hospital stays, with daily information on admissions, in-hospital deaths, demographic characteristics, region, and ICD-10 diagnoses. This allowed us to compare admissions and in-hospital deaths within the same health system and under a unified exposure framework. The cumulative heat metric provides an interpretable measure of sustained exposure and can be translated into absolute excess cases for planning purposes. Sensitivity analyses using alternative exposure windows, humidity-based definitions, primary and secondary diagnoses, and distributed lag non-linear models supported the main mortality findings and confirmed that the mortality signal was concentrated in the first days after heat exposure (see SI).

Several limitations should be considered. First, the study is observational and cannot fully disentangle direct physiological effects of heat from indirect pathways, including changes in care-seeking, admission practices, or hospital operations. Second, the analysis is limited to inpatient events. Out-of-hospital deaths, emergency department visits without admission, ambulatory care, and long-term care are not included, so our estimates do not capture the full health burden of heat. Third, exposure assignment was based on regional temperature averages and cannot account for individual-level exposure, housing conditions, occupational exposure, air conditioning, or behavioral adaptation. Fourth, air pollution was not included because suitable data were not available. However, comparable nationwide evidence suggests that heat-morbidity estimates are often only modestly affected by adjustment for air pollutants [17]. Finally, diagnoses are based on administrative coding and may be affected by documentation practices, especially during periods of health-system strain.

### Policy implications

Our findings suggest that heat-health preparedness should account for cumulative exposure rather than relying only on single-day temperature thresholds. Sustained heat was associated with clear increases in in-hospital mortality, especially among older adults and patients with respiratory, infectious, and endocrine or metabolic conditions. We find an increase in risk both for already admitted before a heat period, bearing important implications for adaptation measures potentially needed within the health sector itself, and increased risk on the day of admission which may reflect the need for improved early warning systems and ways to identify vulnerable population groups earlier. At the same time, admissions did not rise uniformly, indicating that hospital demand during heat periods cannot be inferred from mortality risk alone.

Preparedness plans should therefore combine heat forecasts with information on age, diagnosis, and regional vulnerability. Hospitals may need targeted protocols for high-risk inpatients during prolonged heat periods, while community and primary-care services may be important for earlier identification and support of vulnerable people before hospital admission. Translating relative risks into absolute excess cases can support capacity planning, future burden estimation, and prioritization of climate adaptation measures.

## Conclusion

Sustained heat exposure is associated with a substantial increase in in-hospital mortality in Austria, particularly among older adults and patients with respiratory, infectious, and endocrine or metabolic conditions. Hospital admissions show a more selective pattern, declining overall but increasing for specific heat-sensitive diagnoses. These findings show that mortality and admissions provide different signals of heat-related health burden. Heat-health preparedness should therefore account for cumulative exposure and target vulnerable diagnostic and demographic groups, rather than assuming a uniform increase in hospital demand during heatwaves.

## CRediT authorship contribution statement

Katharina Ledebur: Methodology, Formal analysis, Software, Visualization, Writing – original draft, Writing – review and editing.

Katharina Brugger: Interpretation, Writing – review and editing. Andrea Schmidt: Interpretation, Writing – review and editing.

Marianne Bügelmayer-Blaschek: Interpretation, Writing – review and editing. Martin Schneider: Interpretation, Writing – review and editing.

Andrea Hochebner: Interpretation, Writing – review and editing.

Peter Klimek: Conceptualization, Methodology, Supervision, Funding acquisition, Writing – review and editing.

## Data availability

Meteorological data constitute the intellectual property of UBIMET, are legally protected, and may not be disclosed to third parties. Data on hospital admissions and deaths involve individual health information and cannot be shared. Aggregated formats can be provided upon reasonable request.

## Ethics approval

This study used pseudonymized administrative hospital records. The ethics committee of the Medical University of Vienna approved the study [1768/2025 HeatProtect - Entwicklung und datenbasierte Evaluation klimafreundlicher Maßnahmen zur Minderung hitzebedingter Gesundheitsrisiken - eine retrospektive Datenanalyse]. All analyses complied with applicable data protection regulations.

## Acknowledgments

We thank UBIMET for providing the meteorological data used in this study.

## Appendix Supplementary Methods

### Data and study population

The hospital dataset represents claims data of inpatient stays in Austria during the study period (2007– 2019). Each record corresponds to one hospital stay and includes admission and discharge dates, age group, sex, region of residence, region of care, diagnoses (primary and secondary), and discharge status.

For admission analyses, temperature exposure was assigned based on the patient’s region of residence. For mortality analyses, exposure was assigned based on the region of care. Deaths occurring after long hospital stays were linked to temperature exposure at the day of death (discharge date).

### Exposure definition

Daily temperature data were aggregated to care regions using population-weighted averages. A binary heat-day indicator was defined for each region *r* and day *t* based on whether daily maximum temperature exceeded a threshold *τ* (26–32*^◦^*C).

Cumulative exposure was defined as a rolling count of heat days over the preceding 14 days:

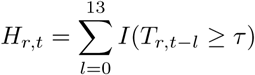

Thus, *H_r,t_*represents the number of heat days within the previous 14-day window, including the current day.

In additional specifications, exposure was restricted to periods with at least *Z* consecutive heat days within the same window (*Z* = 1–5).

### Non-linear exposure transformation

To allow for non-linear effects, cumulative exposure was transformed as

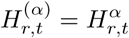

where *α >* 0 controls the shape of the exposure–response relationship, how cumulative heat exposure translates into risk. Values *α* = 1 correspond to a linear effect, values *α <* 1 imply diminishing marginal effects of additional heat days (saturation), whereas *α >* 1 implies increasing marginal effects (accumulation or compounding risk). This parametrization provides a parsimonious approximation of monotonic non-linear exposure–response relationships.

### Statistical model

Associations were estimated using Poisson regression models with robust standard errors:

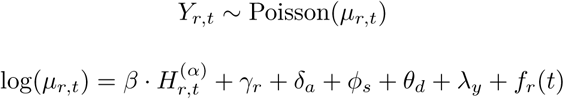

where *Y_r,t_*denotes daily counts of admissions or deaths, *γ_r_* are region fixed effects, *δ_a_* age group indicators, *ϕ_s_*sex indicators, *θ_d_*day-of-week indicators, *λ_y_*year fixed effects, and *f_r_*(*t*) a region-specific linear time trend.

The time trend is continuous over the full study period. Year fixed effects account for between-year variation. We only include summer months (June, July, and August).

### Model selection

The parameter *α* was selected using a grid search over predefined values (0.1 to 3). For each heat definition, models were estimated across all candidate values, and the optimal specification was chosen based on the Akaike Information Criterion (AIC), conditional on statistical significance of the exposure term.

### Definition of non-heat periods

Baseline periods were defined as days with no heat exposure in the preceding 14 days, that is *H_r,t_* = 0. Baseline daily counts were computed using data from 2015–2019, stratified by region, age group, and sex.

### Additional cases

For a given exposure level *d*, the incidence rate ratio is given by

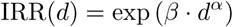

**Figure 6:**
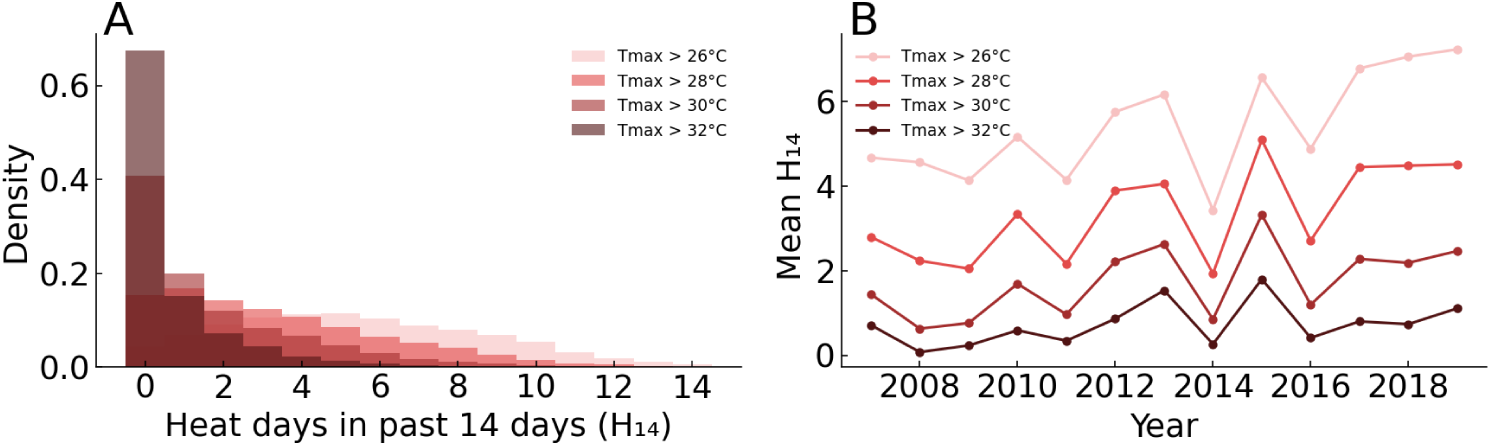
Distribution and temporal variation of cumulative heat exposure. Left panel shows the empirical distribution of *H*_14_ (number of heat days in the preceding 14 days) across all summer region-days (June–August, 2007–2019) for four temperature thresholds. Right panel shows the annual mean *H*_14_ by threshold. Year-to-year variation reflects interannual climate variability, with 2015 representing the highest heat exposure year in the study period.

Additional cases were calculated as

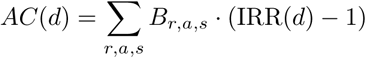

where *B_r,a,s_*denotes the baseline daily count for each region, age group, and sex stratum.

Additional cases therefore represent expected excess events per day under a given exposure scenario. Daily rates per 100,000 population were calculated using corresponding population denominators. Confidence intervals were derived from the standard errors of *β*.

### Interpretation of IRR_5_*_d_* and IRR_14_*_d_*

IRR_5*d*_ and IRR_14*d*_ were obtained by evaluating the model at *H* = 5 and *H* = 14, respectively:

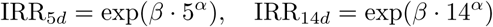

These values represent model-based exposure scenarios rather than averages over observed days.

#### Sensitivity analysis for exposure and outcome definitions

**Figure 7:**
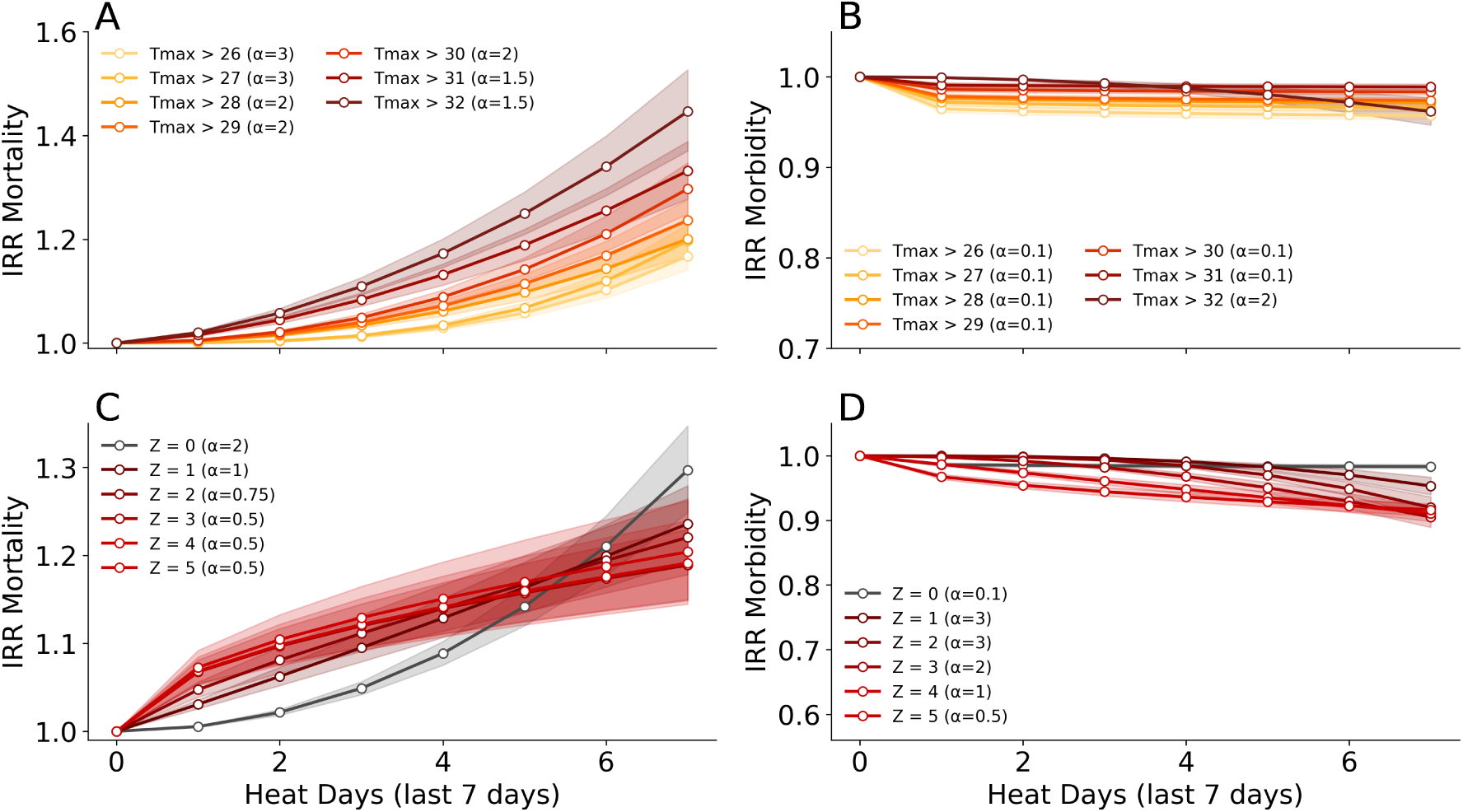
Impact of number of heat days (daily maximum temperature above a threshold) in the last 7 days on in-hospital mortality and admission across different temperature thresholds and persistence definitions. Incidence Rate Ratios (IRRs) for mortality (A) and admission (B) by threshold of daily maximum temperature (Tmax > 26°C to Tmax > 32°C). Higher thresholds are associated with stronger mortality effects. Admission effects remain weak or negative at lower thresholds and become more positive only at the highest thresholds. IRRs for mortality (C) and admission (D) by number of prior consecutive hot days. Shaded bands represent 95% confidence intervals. *α* denotes the scaling parameter optimized for model fit. Only statistically significant estimates are shown.

**Figure 8:**
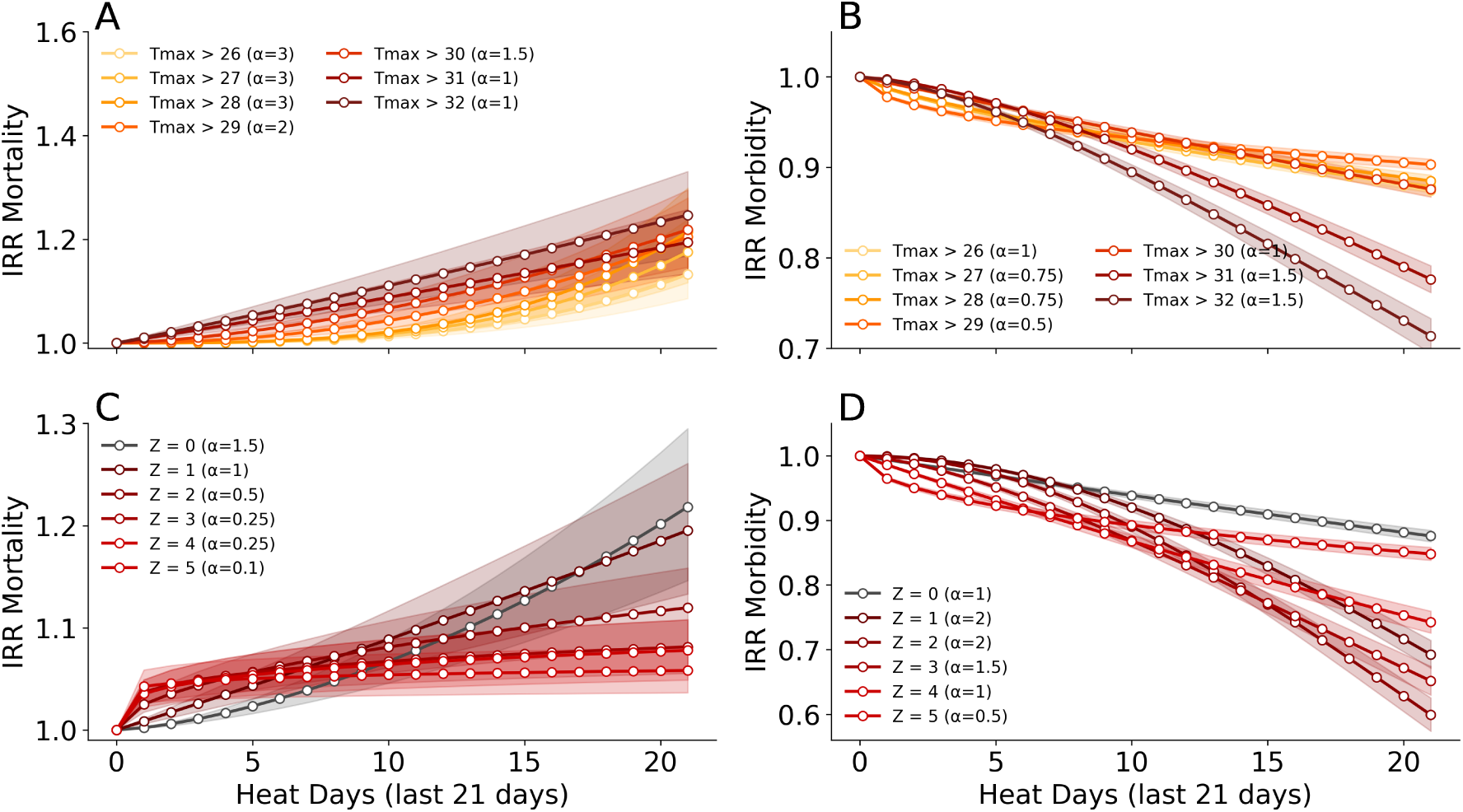
Impact of number of heat days (daily maximum temperature above a threshold) in the last 21 days on in-hospital mortality and admission across different temperature thresholds and persistence definitions. Incidence Rate Ratios (IRRs) for mortality (A) and admission (B) by threshold of daily maximum temperature (Tmax > 26°C to Tmax > 32°C). Higher thresholds are associated with stronger mortality effects. Admission effects remain weak or negative at lower thresholds and become more positive only at the highest thresholds. IRRs for mortality (C) and admission (D) by number of prior consecutive hot days. Shaded bands represent 95% confidence intervals. *α* denotes the scaling parameter optimized for model fit. Only statistically significant estimates are shown.

**Figure 9:**
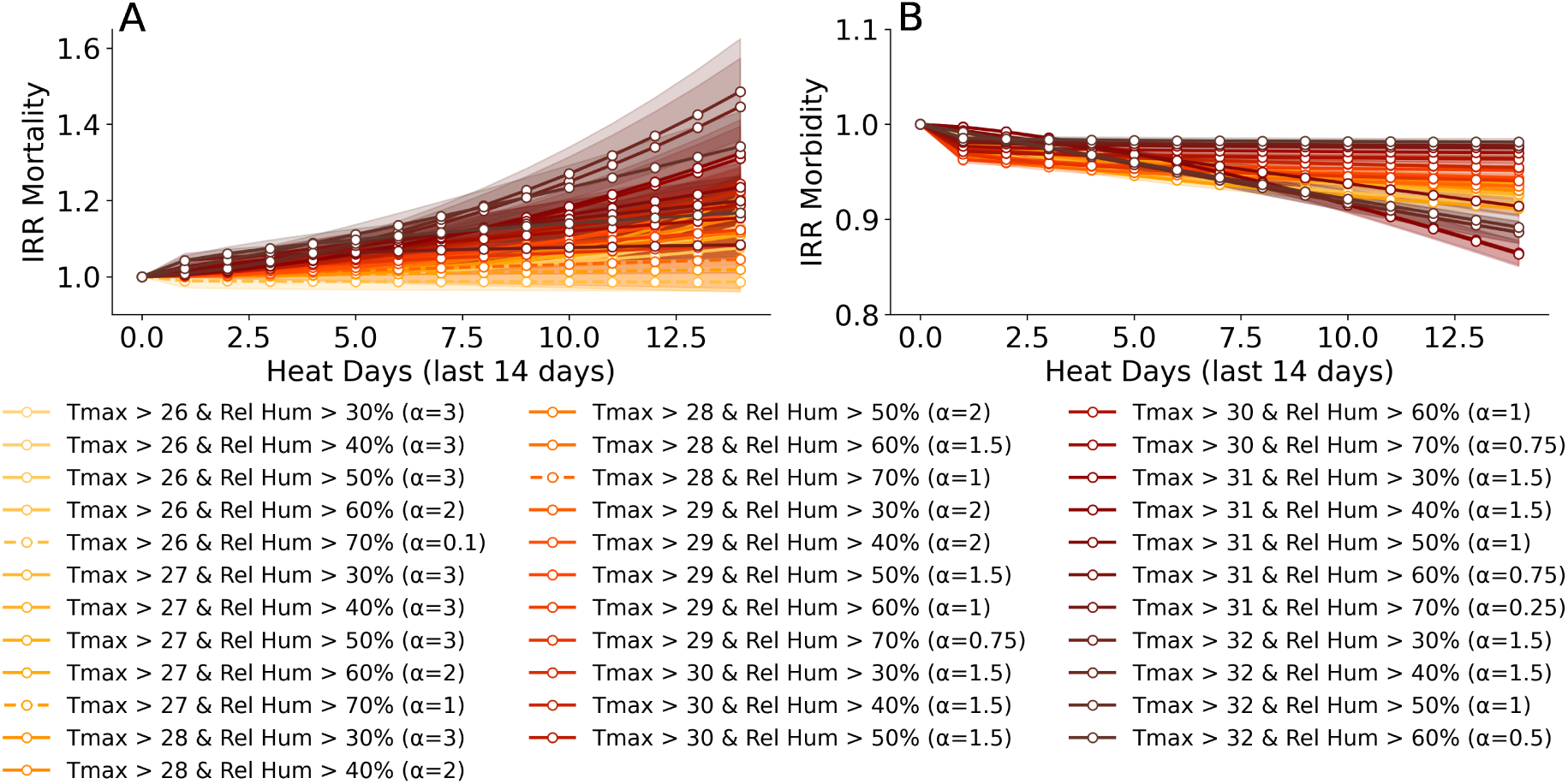
Impact of heat days (daily maximum temperature and relative humidity above a threshold) on mortality and admission across different temperature thresholds and persistence definitions. Incidence Rate Ratios (IRRs) for mortality (A) and admission (B) by threshold of daily maximum temperature (Tmax > 26°C to Tmax > 32°C) and relative humidity (Rel Hum > 30% to Rel Hum > 70%). Higher thresholds are associated with stronger mortality effects. Admission effects remain weak or negative at lower thresholds and become more positive only at the highest thresholds. *α* denotes the scaling parameter optimized for model fit. Only statistically significant estimates are shown.

**Figure 10:**
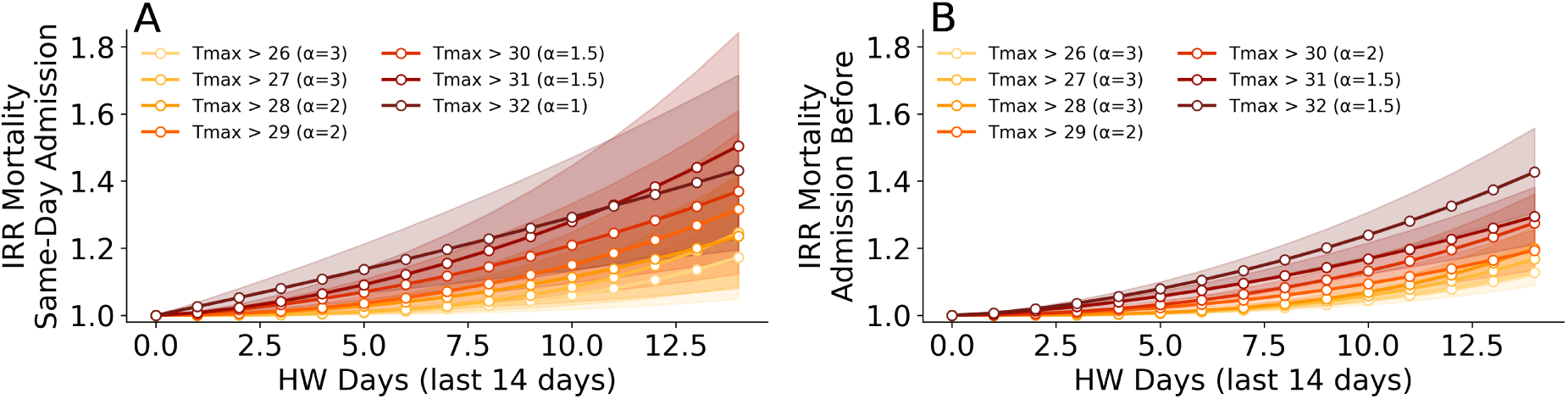
Impact of heat days on on in-hospital mortality by timing of admission. Incidence Rate Ratios (IRRs) for mortality among patients admitted on the same day (A), and mortality among patients who were admitted before the day of death (B) by threshold of daily maximum temperature (Tmax > 26°C to Tmax > 32°C). *α* denotes the scaling parameter optimized for model fit. Only statistically significant estimates are shown.

#### Sensitivity analysis using distributed lag non-linear models (DLNMs)

To assess the robustness of our findings and evaluate the lag structure of heat effects on in-hospital mortality and hospital admissions, we fit DLNMs as a sensitivity analysis. DLNMs simultaneously model the exposure-response and lag-response dimensions through a cross-basis of spline functions, and represent the standard methodological framework in the environmental epidemiology literature on temperature and health [4], [23], [24], [25]. We aggregate daily death and admission counts across all age groups and sexes to the region-day level. For each Austrian care region, we fit a separate Poisson GLM with a cross-basis for daily maximum temperature as the primary exposure, controlling for day of week, year fixed effects, and a within-summer time trend. The cross-basis was constructed with a natural cubic spline in the exposure dimension (df=4) and log-spaced interior knots at lags 1, 3, and 8 for the lag dimension (maximum lag 21 days), following Gasparrini *et al.* (2013) [23]. Region-specific coefficients and variance-covariance matrices were pooled using inverse-variance weighting. All models were estimated with HC0 heteroskedasticity-robust standard errors. Predictions were centered at 20°C.

**Figure 11:**
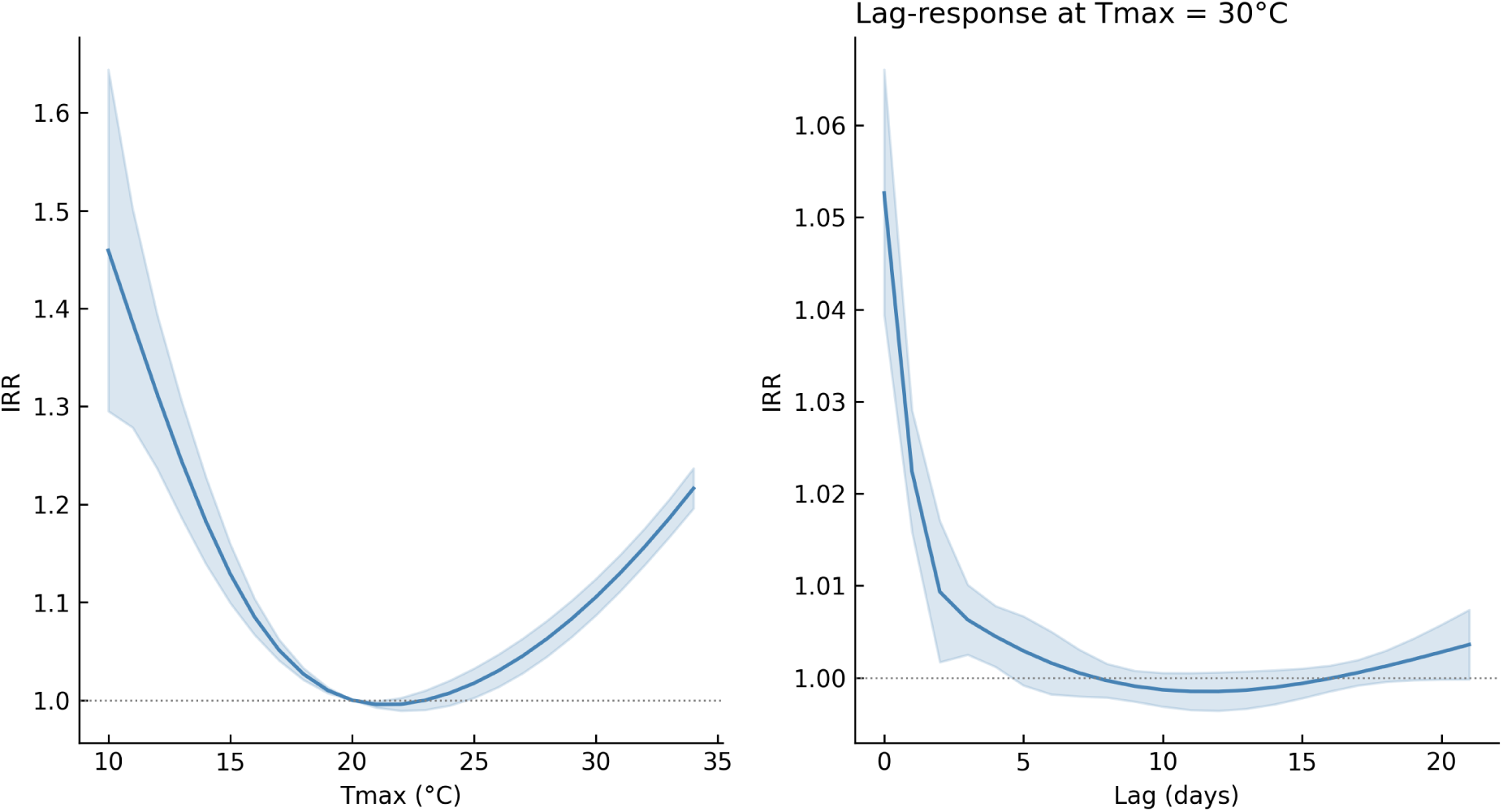
Distributed lag non-linear model results for in-hospital mortality. Left panel shows the cumulative exposure-response (IRR relative to 20°C), right panel shows the lag-response at 30°C. The cross-basis was constructed with a natural cubic spline (df=4) in the exposure dimension and log-spaced interior knots at lags 1, 3, and 8 in the lag dimension (maximum lag 21 days). Region-specific estimates were pooled using inverse-variance weighting across all Austrian care regions.

**Figure 12:**
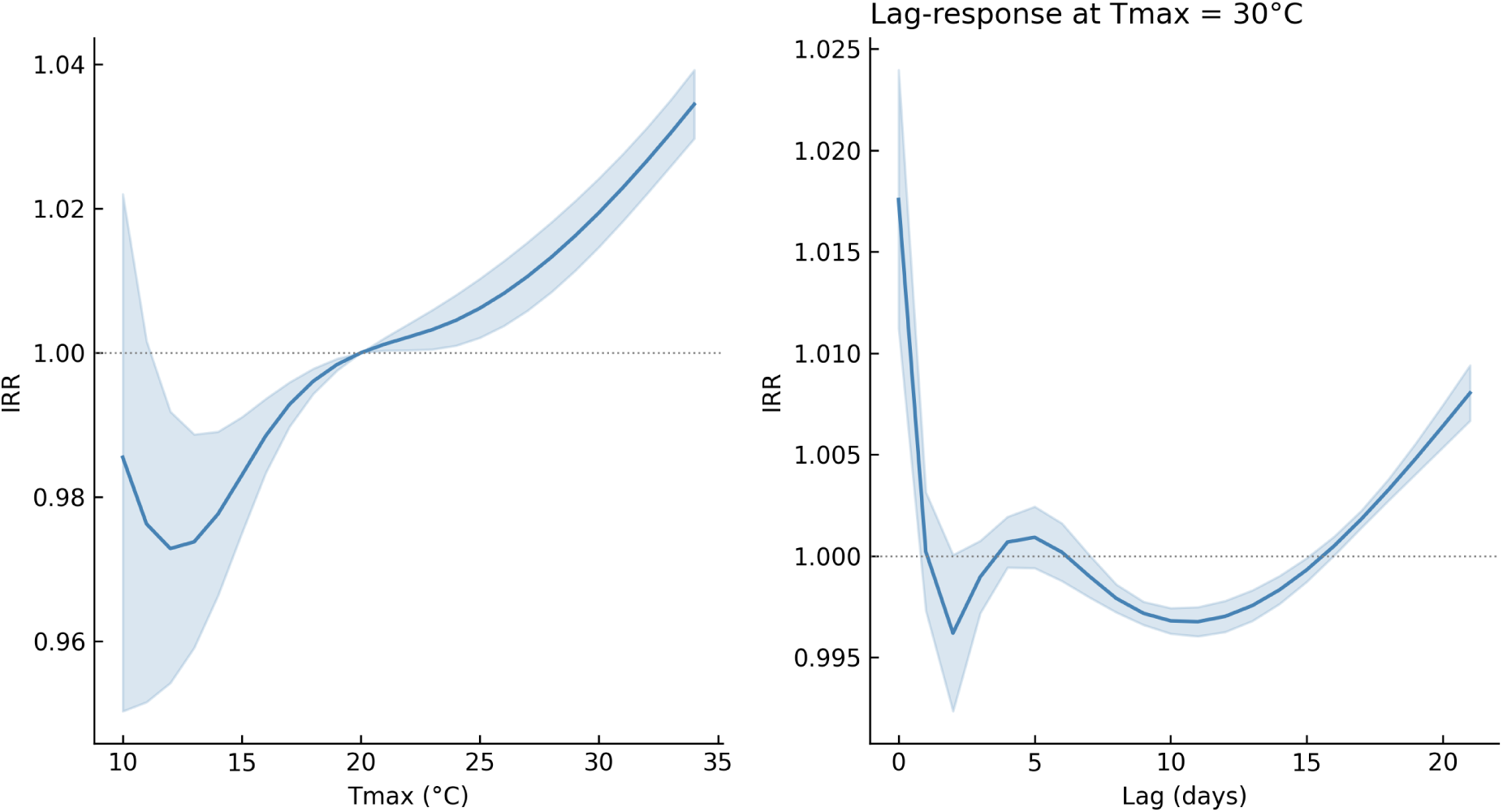
Distributed lag non-linear model results for hospital admissions. Left panel shows the cumulative exposure-response (IRR relative to 20°C), right panel shows the lag-response at 30°C. Specification as in Figure 11.

The cumulative exposure-response for mortality (Figure 11) shows a monotonically increasing association above approximately 22°C, with a cumulative IRR of 1*·*101 (95% CI 1*·*077–1*·*127) at 30°C relative to 20°C. This is directionally consistent with our primary alpha-model estimates. The lag-response at 30°C reveals that the mortality effect is strongly concentrated at lag 0–2, declining sharply thereafter and remaining close to null beyond lag 5. This concentration of the acute signal in the first two days provides empirical support for our primary modelling choice. The rolling accumulation window captures the dominant portion of the heat-mortality signal, and longer uniform weighting across 14 days does not materially misrepresent the underlying lag structure. For hospital admissions, the cumulative exposure-response (Figure 12) shows a negative association at temperatures below approximately 22°C and a modest positive association above 30°C (IRR 1*·*019, 95% CI 1*·*015–1*·*024). This differs from the negative overall admissions effect in our primary analysis, which compares days with accumulated heat exposure to days with zero qualifying heat days. The DLNM instead compares continuously varying daily temperatures to a 20°C reference, capturing a different contrast. The positive signal at extreme temperatures in the DLNM is consistent with acute heat-related presentations, while the overall negative effect in our primary analysis likely reflects broader behavioral responses. The lag-response for admissions mirrors the mortality pattern, with effects concentrated at lag 0–1.

A concern with DLNMs is sensitivity to the number and placement of spline knots, which are typically chosen by convention rather than estimated from data. Table 2 shows the cumulative IRR at 30°C across all combinations of lag knot specification (log-spaced with 2, 3, or 4 interior knots) and exposure dimension degrees of freedom (df = 3–6). For df = 3 and df = 4, estimates are stable across all lag specifications, with mortality IRR at 30°C ranging from 1*·*106 to 1*·*113 (95% CI approximately 1*·*086–1*·*142) and admissions IRR ranging from 1*·*018 to 1*·*023 (95% CI approximately 1*·*015–1*·*027). Specifications with df = 5 and df = 6 produce degenerate estimates, indicating numerical failure in the cross-basis construction or the pooling step. These failures occur despite the use of nationwide data at daily resolution covering thirteen summers. The stable df = 3 and df = 4 specifications yield cumulative IRR estimates directionally consistent with our primary results, supporting the robustness of the main findings to the choice of modelling framework.

**Table 2:** Cumulative IRR at 30°C relative to 20°C across DLNM specifications. Lag knot specifications use log-spaced interior knots: log2 (lags 1, 8), log3 (lags 1, 3, 8), log4 (lags 1, 3, 6, 8). df refers to degrees of freedom in the exposure dimension. Degenerate estimates (*^†^*) indicate numerical failure in the cross-basis construction or pooling step.

| Lag spec | df | IRR (95% CI) |  |
| --- | --- | --- | --- |
|  |  | In-hospital mortality | Hospital admissions |
| log2 | 3 | 1.113 (1.087–1.140) | 1.022 (1.017–1.026) |
|  | 4 | 1.106 (1.078–1.135) | 1.018 (1.014–1.023) |
|  | 5 | 4 · 108† | † |
|  | 6 | 0 · 000† | † |
| log3 | 3 | 1.112 (1.086–1.139) | 1.022 (1.018–1.027) |
|  | 4 | 1.107 (1.079–1.135) | 1.019 (1.015–1.024) |
|  | 5 | ~0† | 0 · 000† |
|  | 6 | 0 · 000† | † |
| log4 | 3 | 1.113 (1.087–1.140) | 1.023 (1.018–1.027) |
|  | 4 | 1.113 (1.085–1.142) | 1.020 (1.015–1.025) |
|  | 5 | 3 · 39 × 10<sup>105</sup>† | † |
|  | 6 | † | † |

† Degenerate estimate indicating numerical failure; value not interpretable. Failures occur despite use of nationwide daily data covering thirteen summers, suggesting df ≥ 5 in the exposure dimension is not supported by the data.

#### Analysis for different demographic strata

We additionally evaluate the impact of heat days (Tmax > 30°C) on in-hospital deaths and hospital admissions for different age, sex, and regional strata (Figure 13).

**Figure 13:**
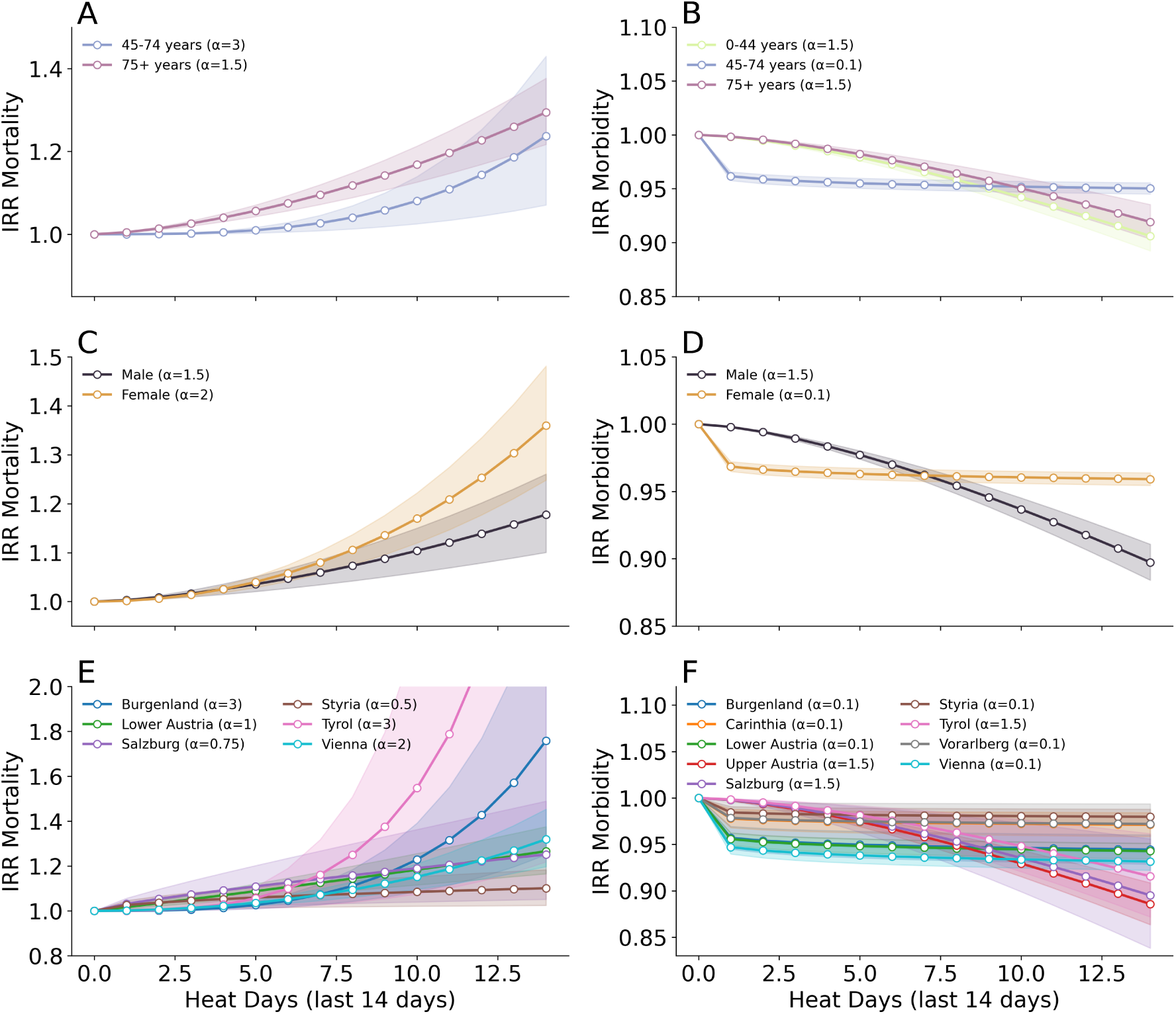
Impact of heat days on mortality and admission across demographic groups and regions. Incidence Rate Ratios (IRRs) for mortality (A) and admission (B) associated with the number of heat days in the past 14 days, stratified by age group (0–44, 45–74, 75+ years). Mortality risk increases steeply with age, while admission shows minimal or negative trends across age groups. IRRs for mortality (C) and admission (D) by sex. Mortality risk increases with heat exposure for both sexes, but more steeply for females; admission effects are negligible or slightly negative. IRRs by region in Austria for mortality (E) and admission (F). Heterogeneity in heat sensitivity is evident across regions, particularly for mortality. Vienna and Tyrol exhibit steeper increases, while several other regions show flatter or no associations, especially for admission. Shaded areas represent 95% confidence intervals.*α* denotes the scaling parameter optimized for model fit. Only statistically significant estimates are shown.

When we stratify by age group, we find significant increases in risk of in-hospital for individuals aged 45 to 74 and 75+ with a maximum of increase in risk of 17.0%(9.0-24.5% 95% CI) and 32.0%(26.6-37.7% 95% CI), respectively (Figure 13 A). For hospital admissions, the association weakens and reverses, with significant decreases by 0.5% (0.1-1% 95% CI) for individuals aged 0 to 44 years and by 2% (1.5-2.5% 95% CI) for individuals aged 45 to 74 years.

When stratifying by sex, we observe a stronger heat-related mortality effect in females, with a maximum increase in risk of 26.5% (21.5–31.7% 95% CI), compared to 22.3% (17.2–27.6% 95% CI) in males (Figure 13B). For hospital admissions, the effect direction diverges: male individuals show a small but significant increase of 1.4% (0.6–2.1% 95% CI), while females exhibit a significant decrease of 1.1% (0.7–1.5% 95% CI). Regional patterns in mortality show the largest relative increases in Burgenland (19.0%, 8.9–29.9% 95% CI), Salzburg (19.1%, 7.5–32.1% 95% CI), and Vienna (20.9%, 15.9–26.2% 95% CI). For hospital admissions, heat-related increases are observed in Burgenland (3.5%, 1.6–5.5% 95% CI), Carinthia (2.8%, 0.6–5.0% 95% CI), Styria (2.6%, 1.3–4.0% 95% CI), and Tyrol (6.6%, 3.0–10.3% 95% CI), while significant reductions appear in Lower Austria (1.0%, 0.3–1.6% 95% CI) and Vienna (1.3%, 0.5–2.0% 95% CI) (Figure 13C).

For all results and absolute numbers of additional in-hospital deaths and hospital admissions see table 3.

**Table 3:**
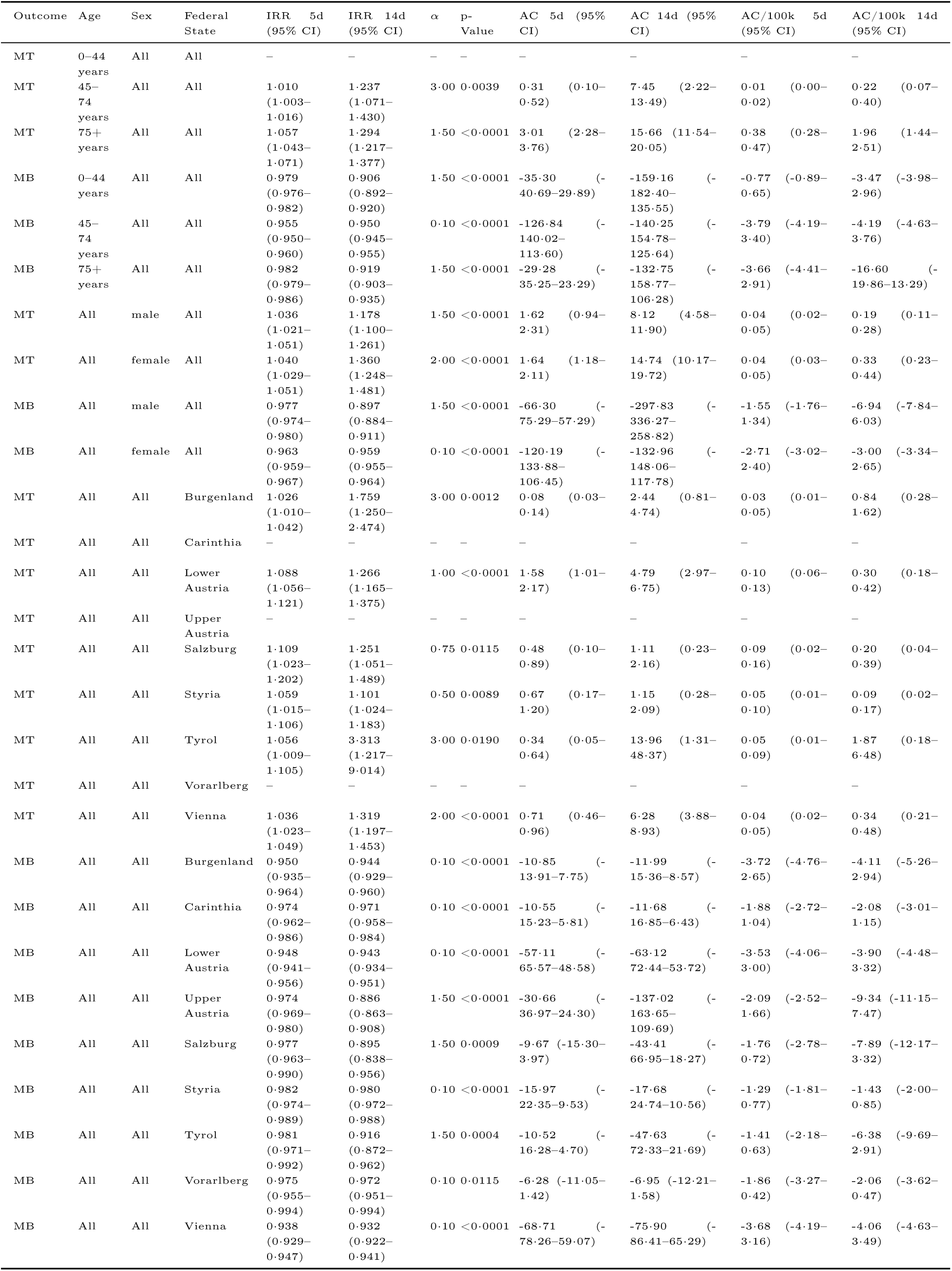
Estimated IRRs and additional cases (AC) and AC per 100,000 with 95% CI across age, sex, and federal state at heat day definition Tmax > 30*◦* C for outcome mortality (MT) and admissions (MB).

#### Analysis using primary diagnoses

**Table 4:**
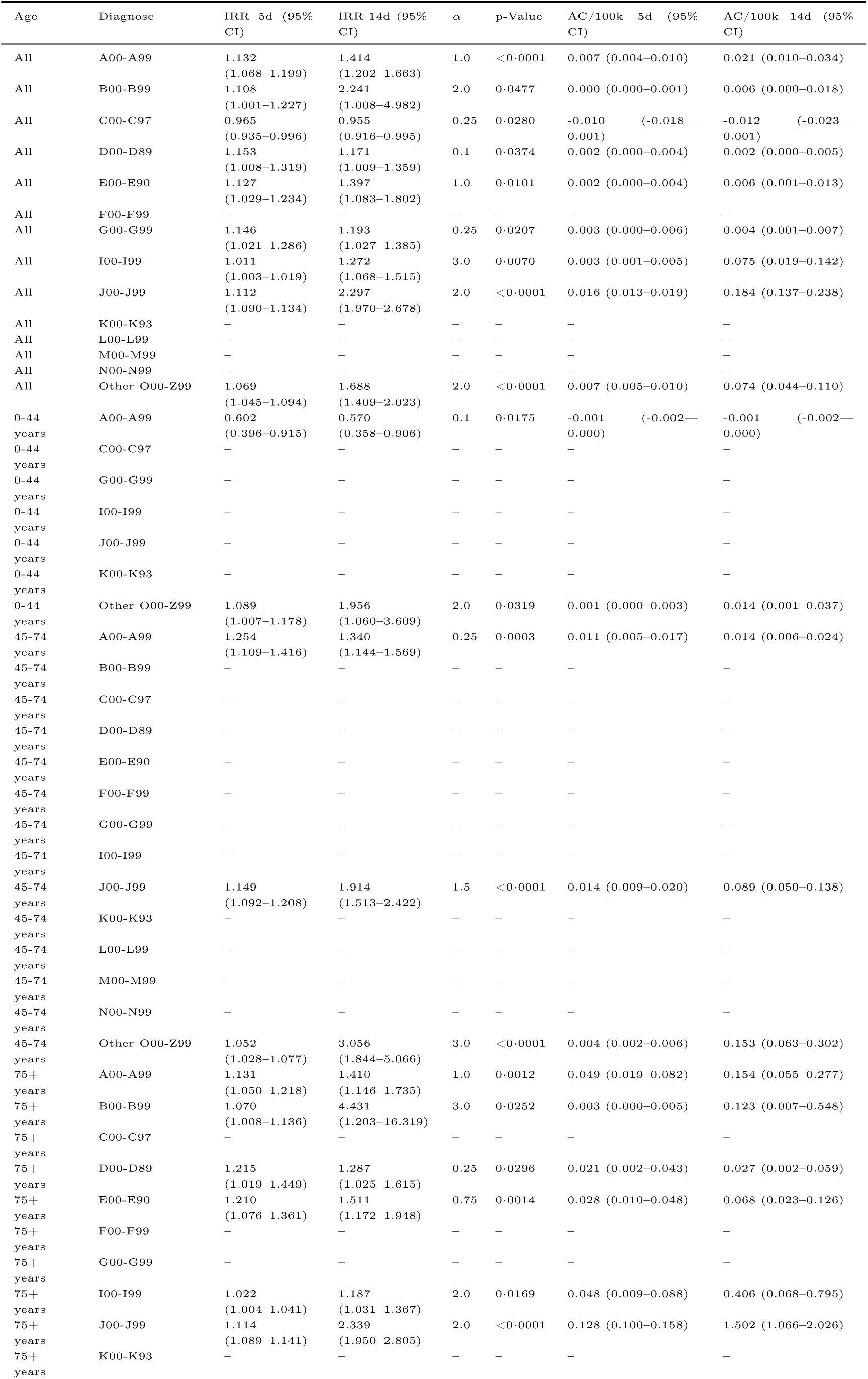
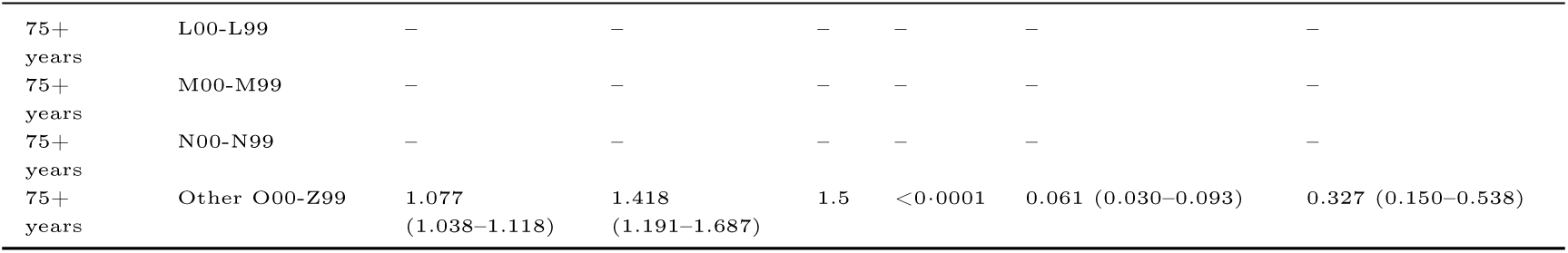
Estimated IRRs and additional cases (AC) per 100k with 95% CI by age group and diagnosis group at heat day definition Tmax > 30C for outcome mortality (MT).

**Table 5:**
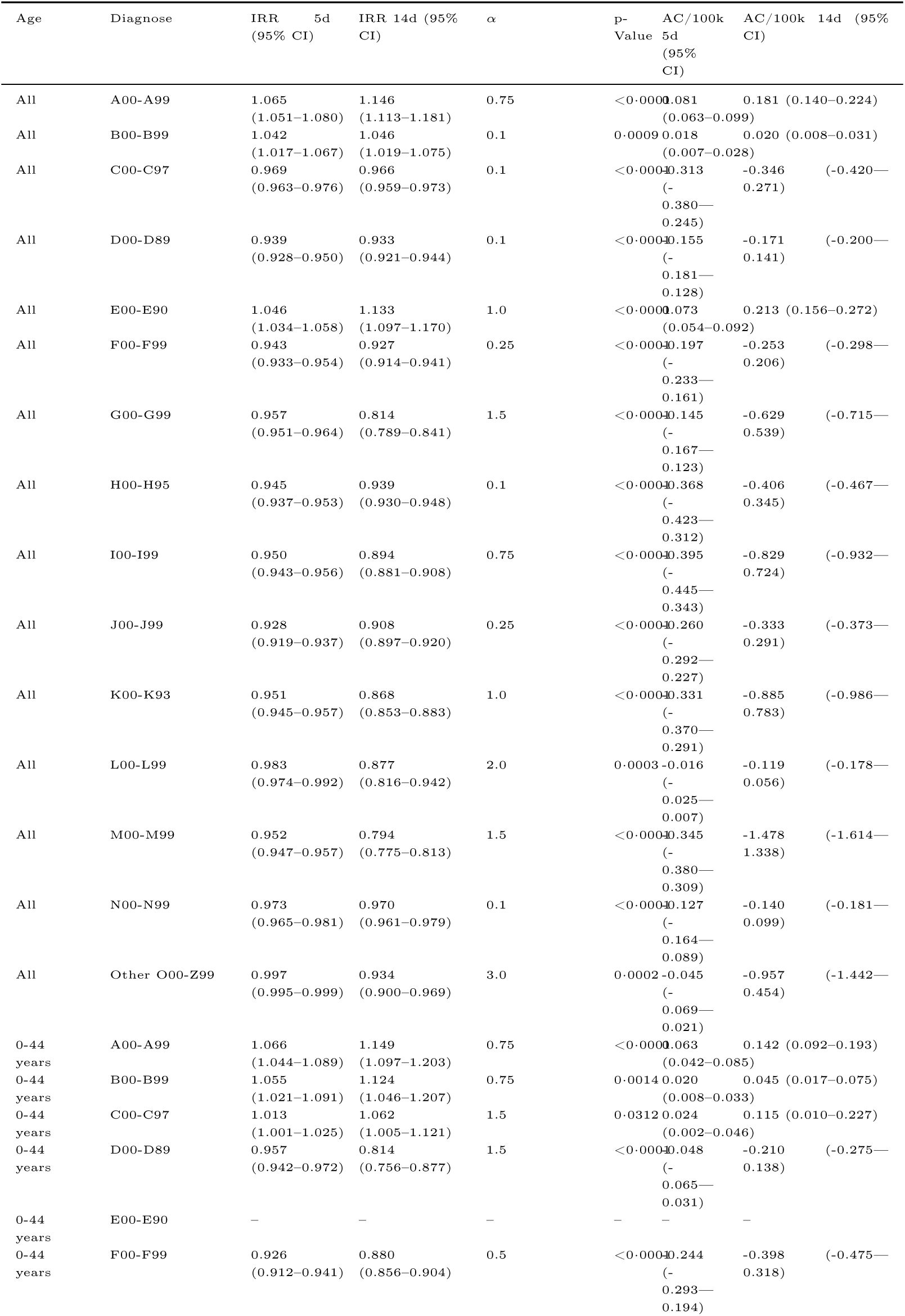
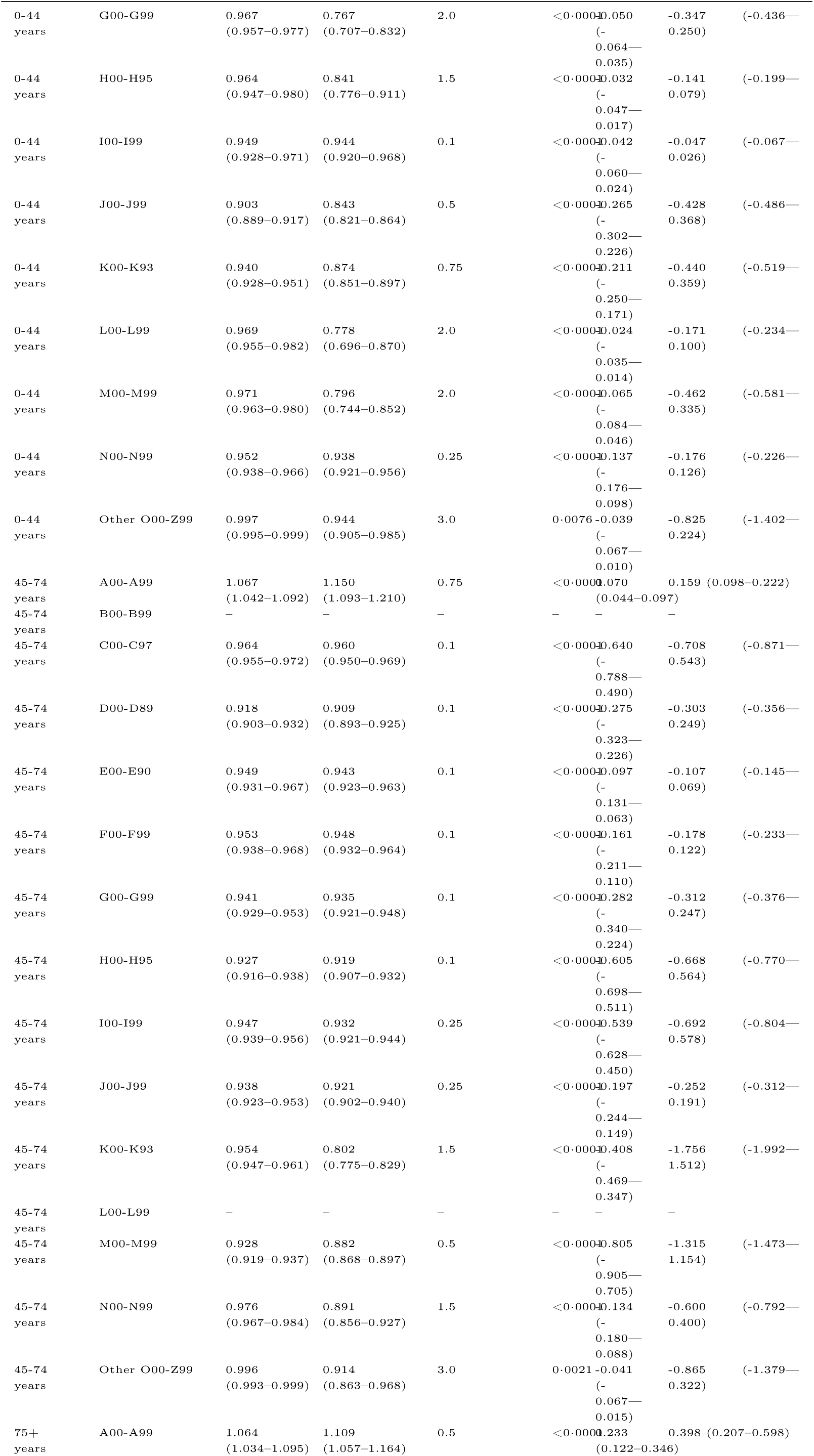
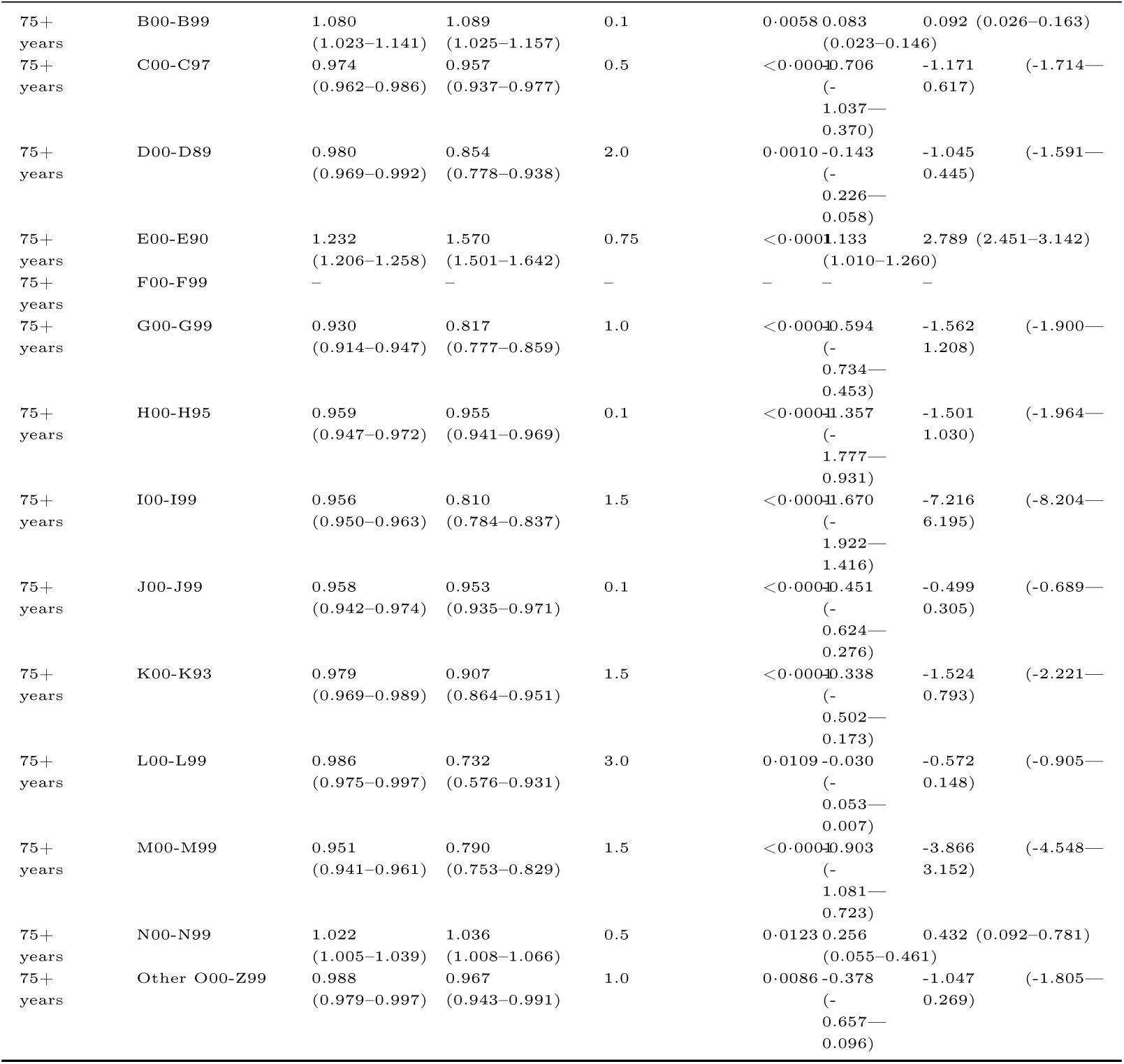
Estimated IRRs and additional cases (AC) per 100k with 95% CI by age group and diagnosis group at heat day definition Tmax > 30C for outcome admissions (MB).

**Table 6:**
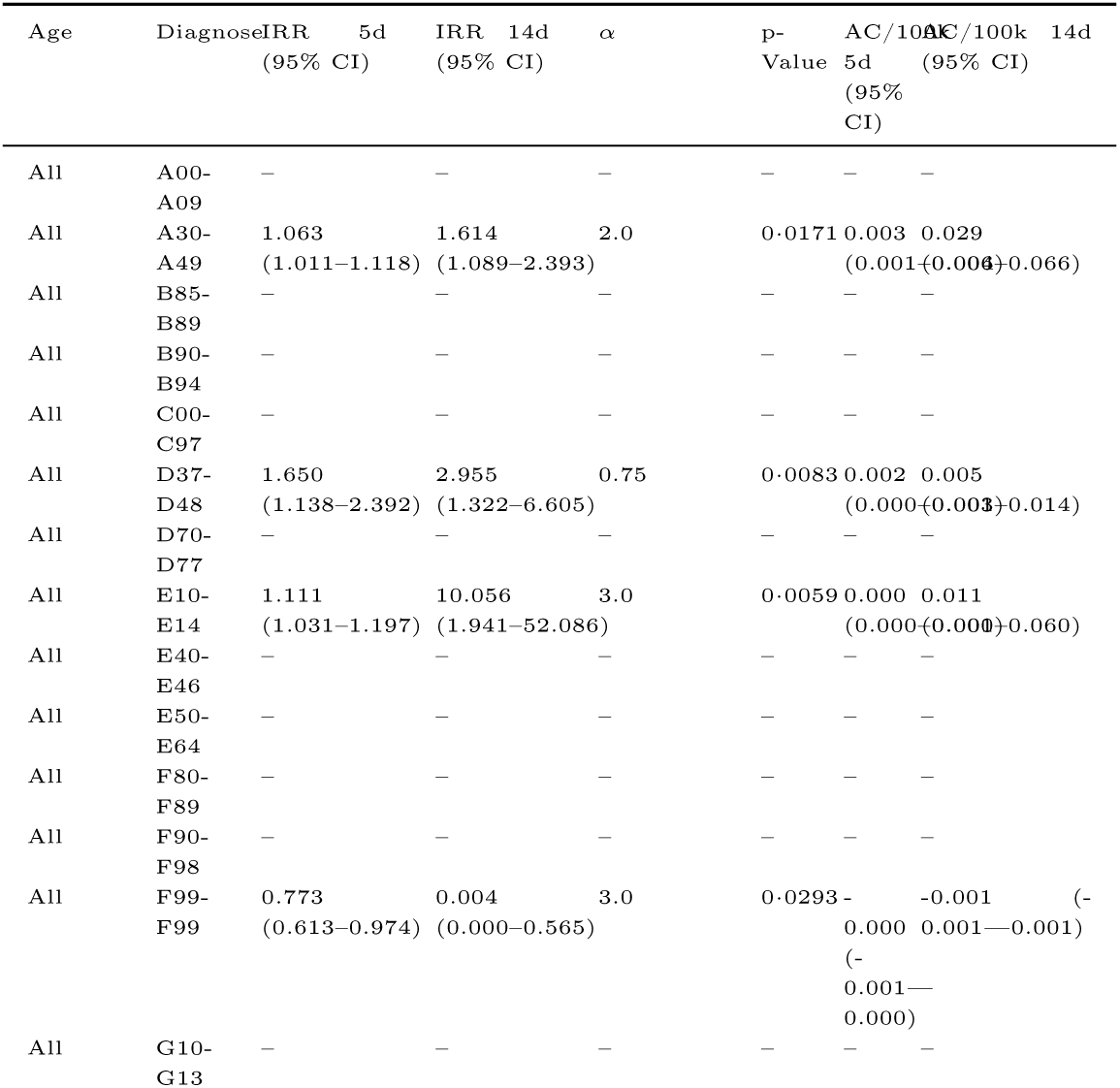
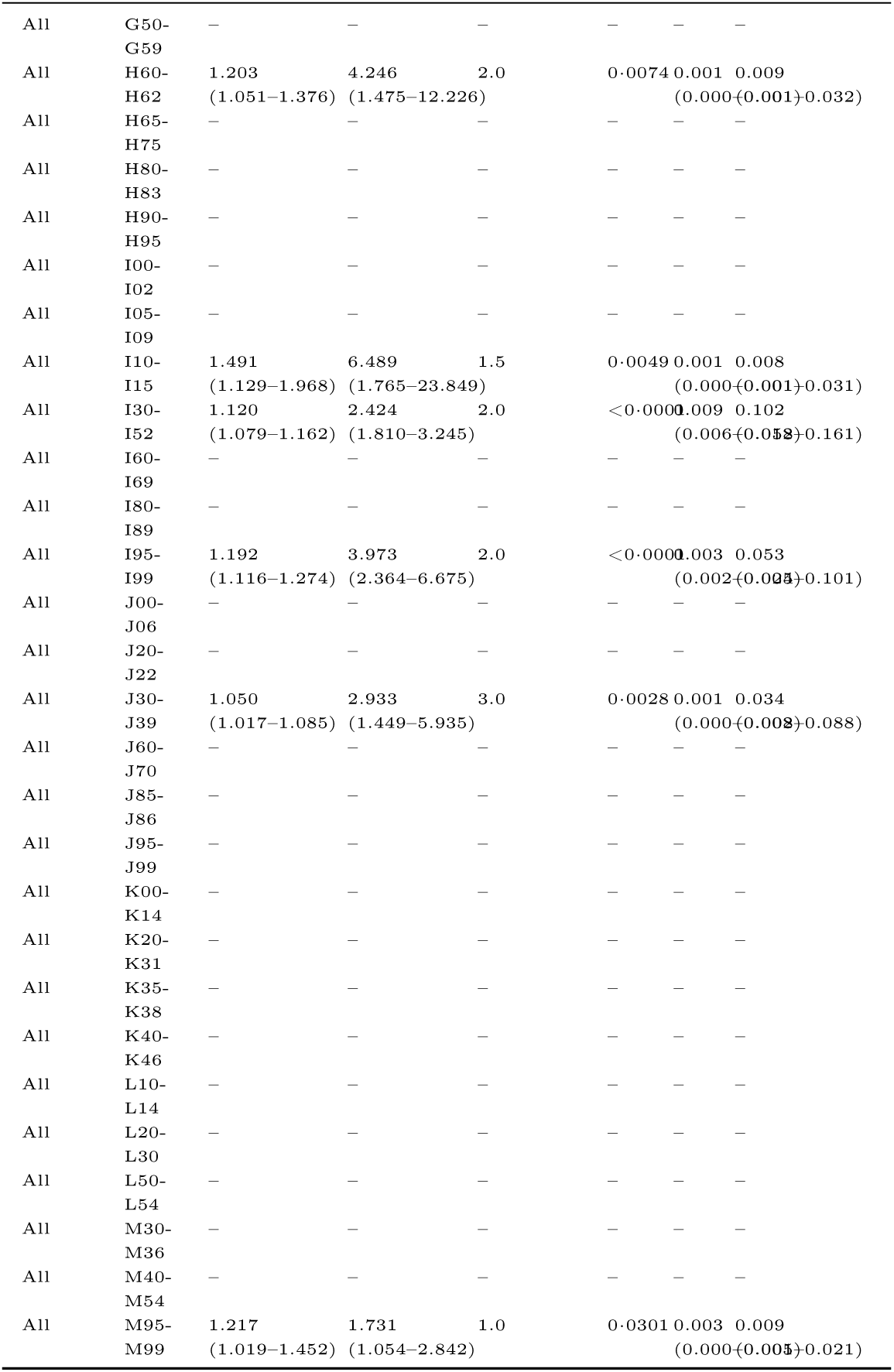
Estimated IRRs and additional cases (AC) per 100k with 95% CI by age group and diagnosis group at heat day definition Tmax > 30C for outcome mortality (MT).

**Table 7:**
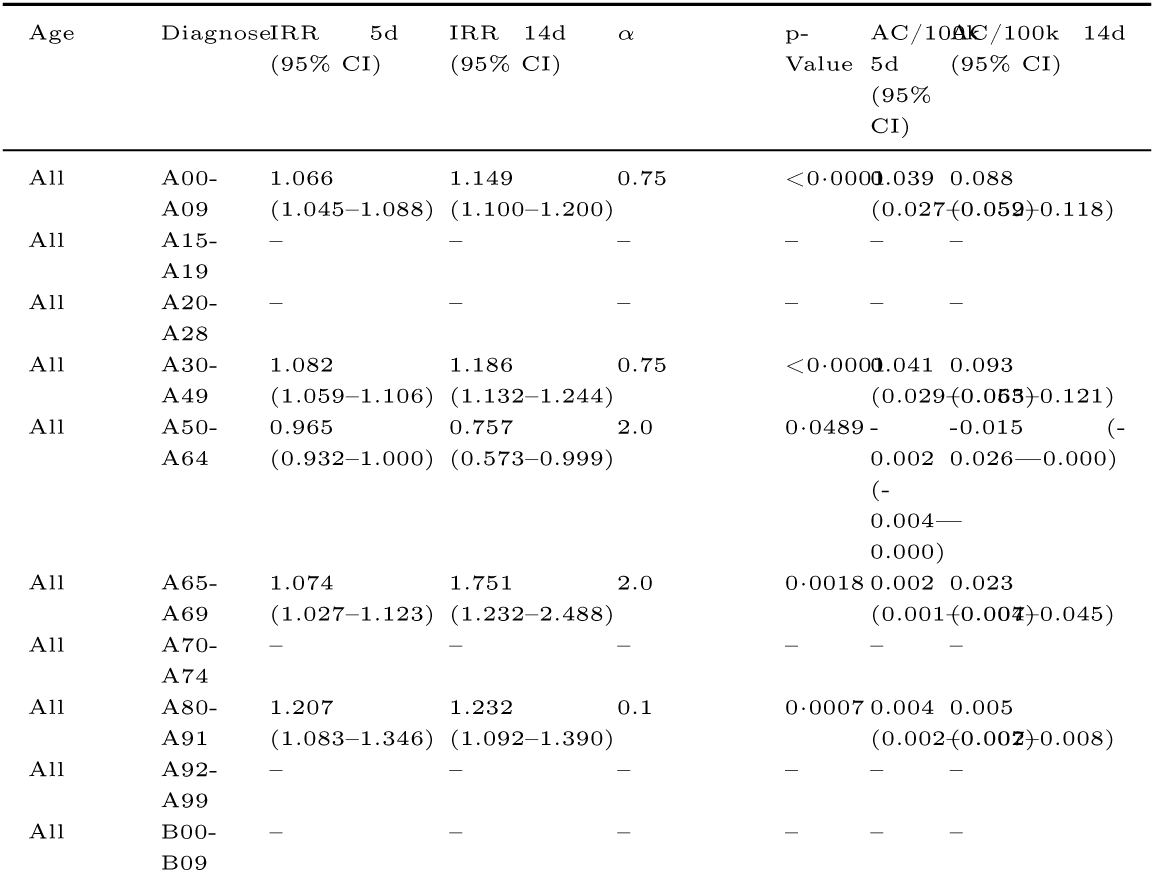
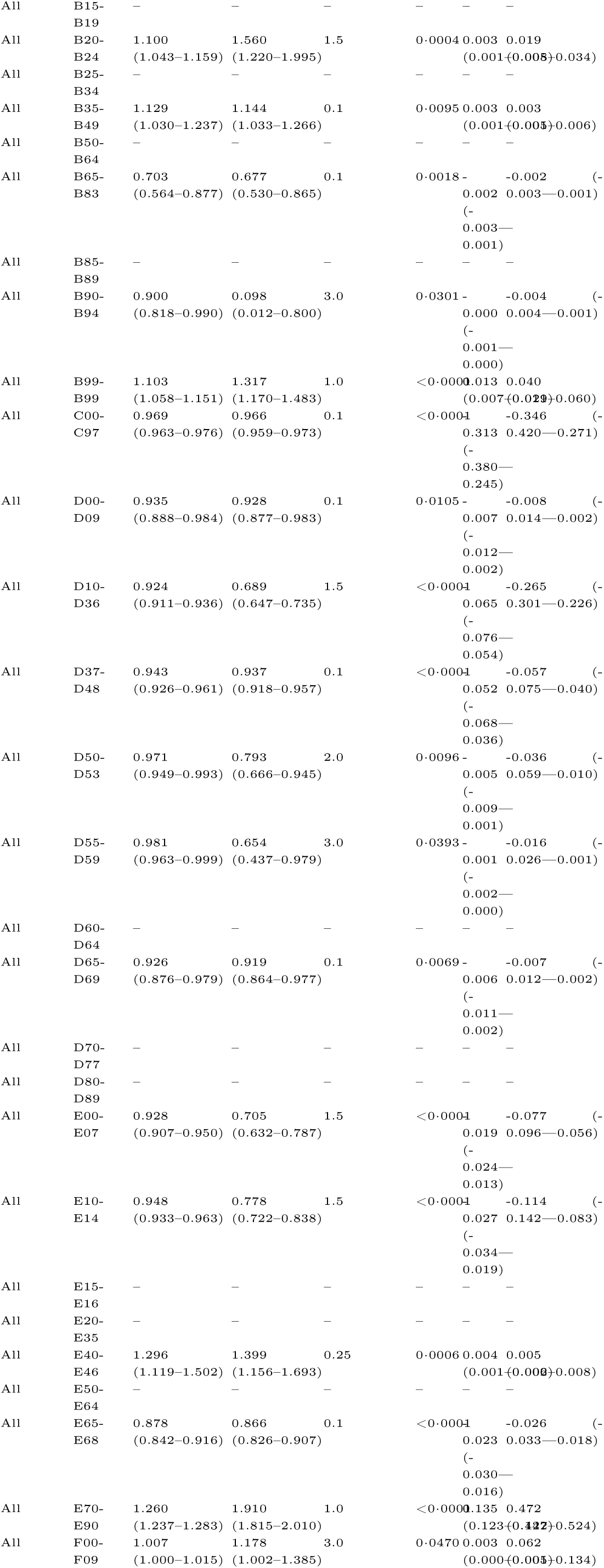
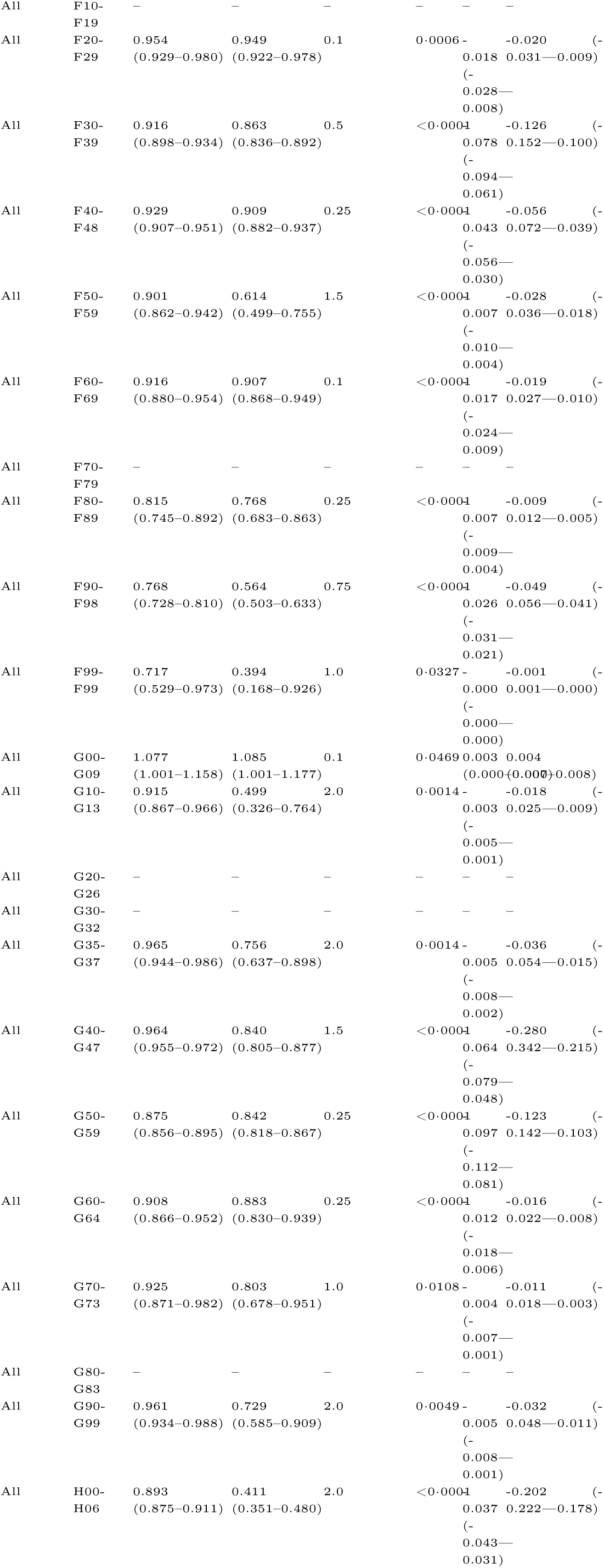
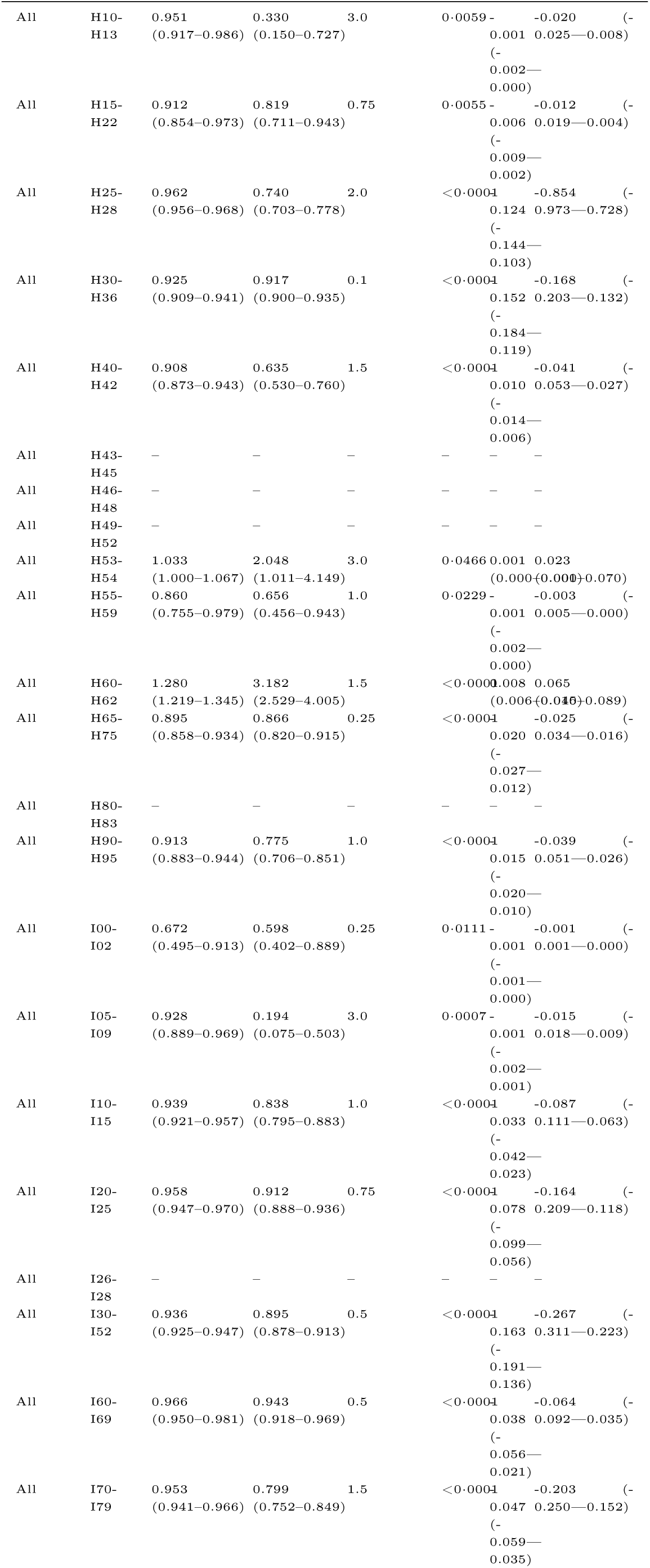
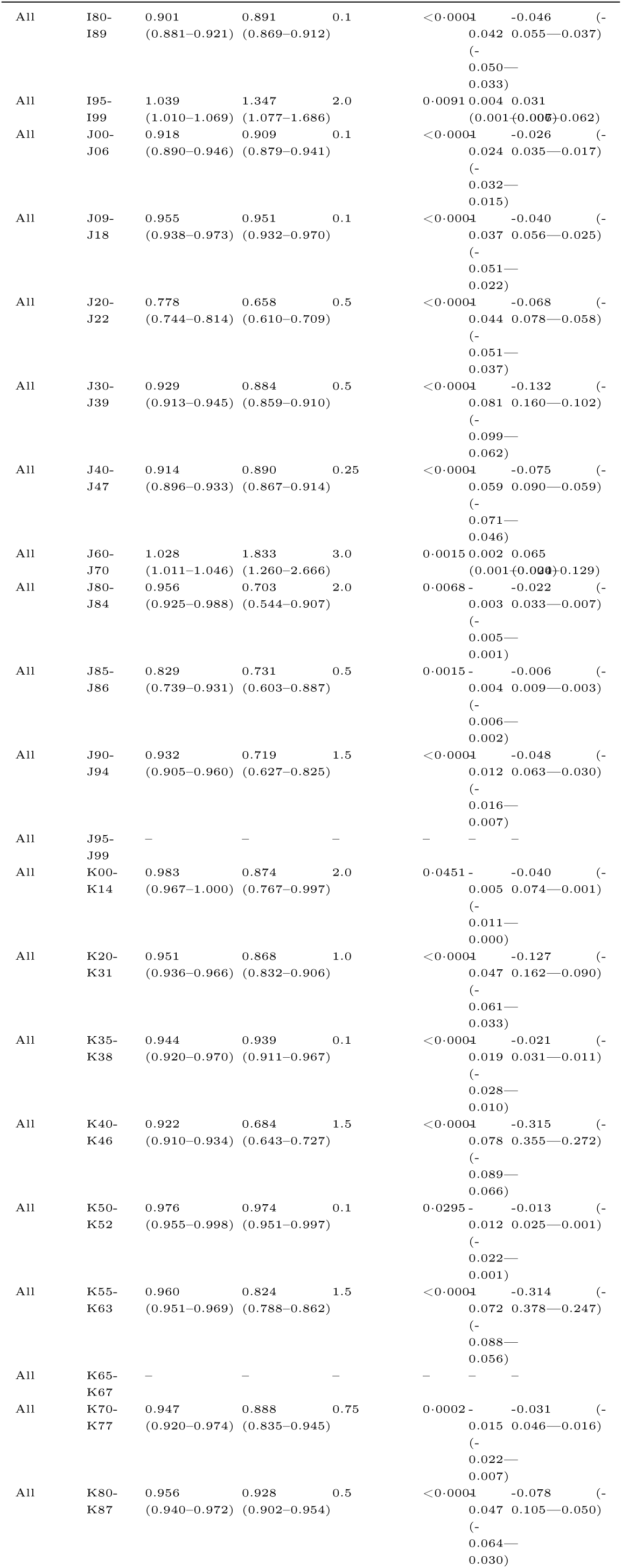
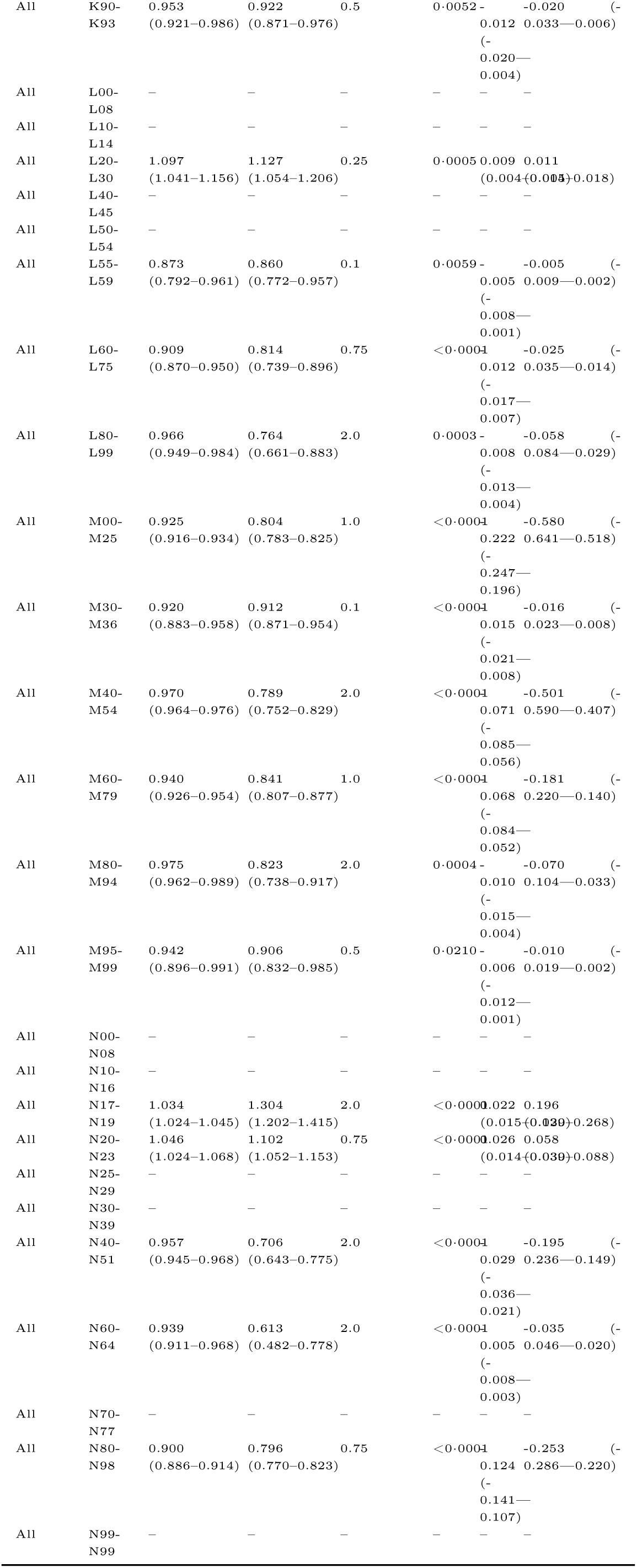
Estimated IRRs and additional cases (AC) per 100k with 95% CI by age group and diagnosis group at heat day definition Tmax > 30°C for outcome admissions (MB).

**Figure 14:**
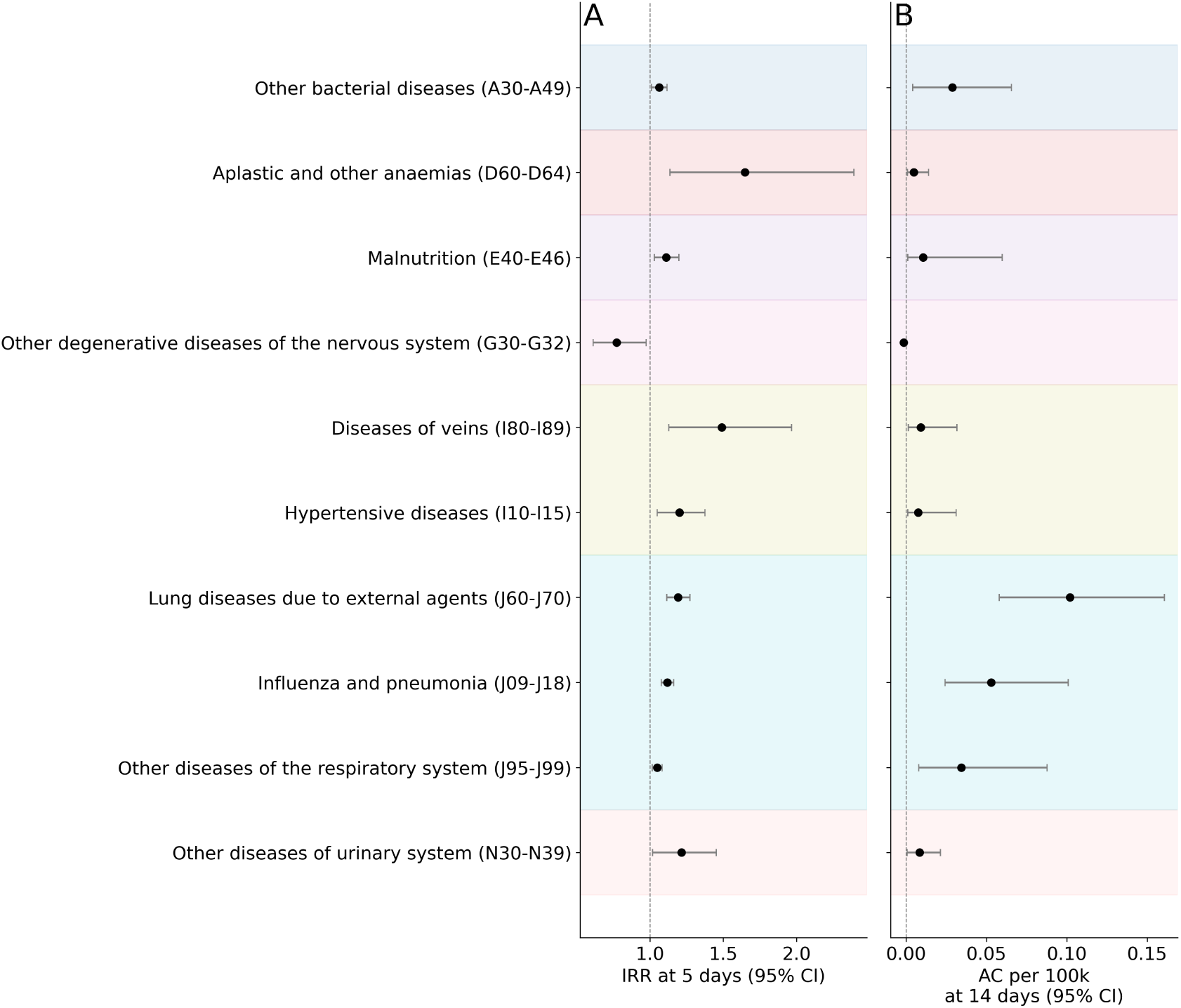
Cumulative effect of heat days on in-hospital death causes. IRR (A) and additional cases (AC) (B) of heat days (Tmax > 30C) across in-hospital death causes after 14 heat days within the past 14 days. We show per ICD-10 chapter (from A00-N99) the three diagnosis groups with the biggest effect. Error bars show the 95% CI. Only statistically significant estimates are shown.

**Figure 15:**
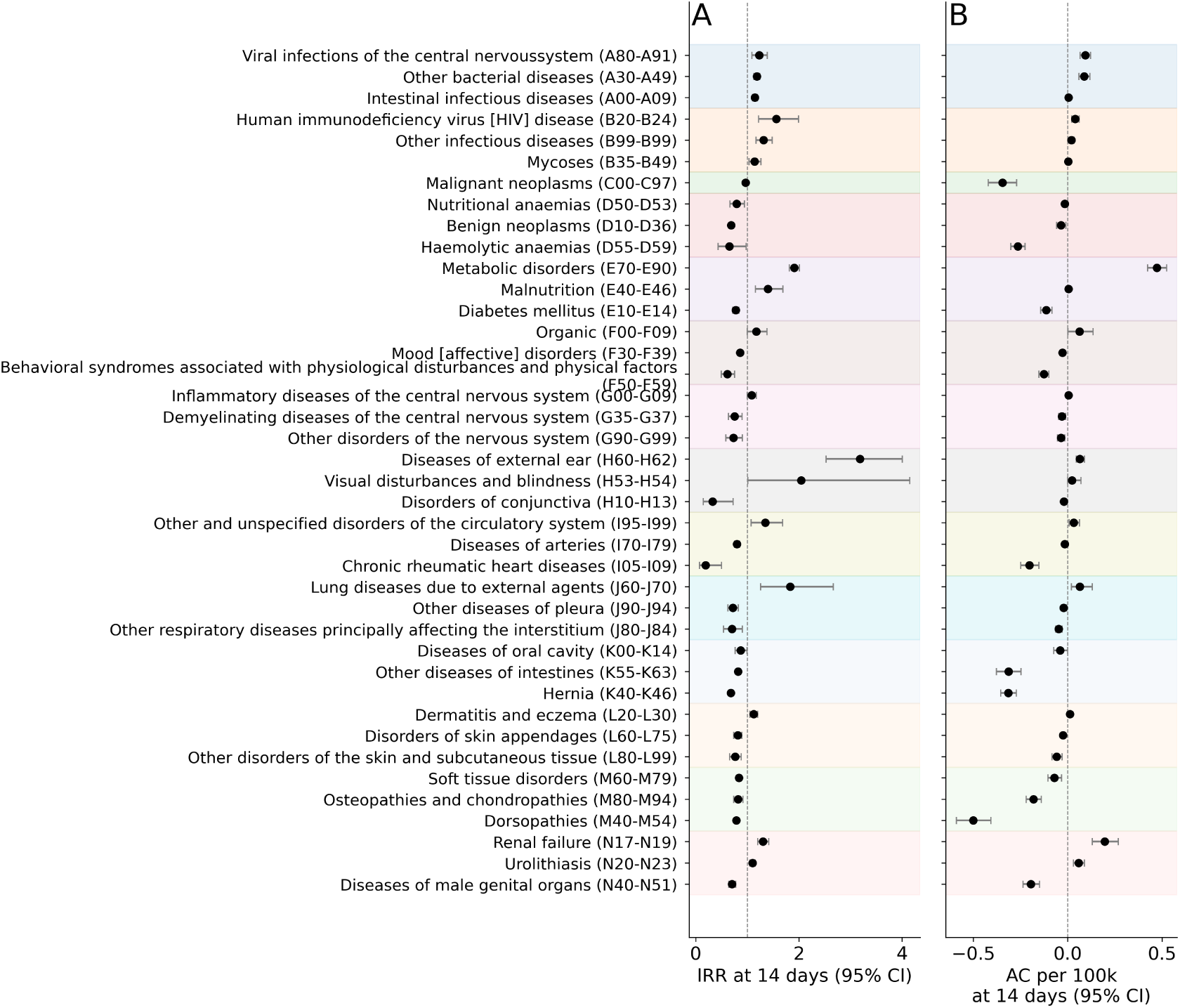
Cumulative effect of heat days on different diagnoses during hospitalization. IRR (A) and additional cases (AC) (B) of heat days (Tmax > 30 C) across diagnosis groups during hospitalization after 14 heat days within the past 14 days. We show per ICD-10 chapter (from A00-N99) the three diagnosis groups with the biggest effect. Error bars show the 95% CI. Only statistically significant estimates are shown.

#### Analysis using primary and secondary diagnoses

**Figure 16:**
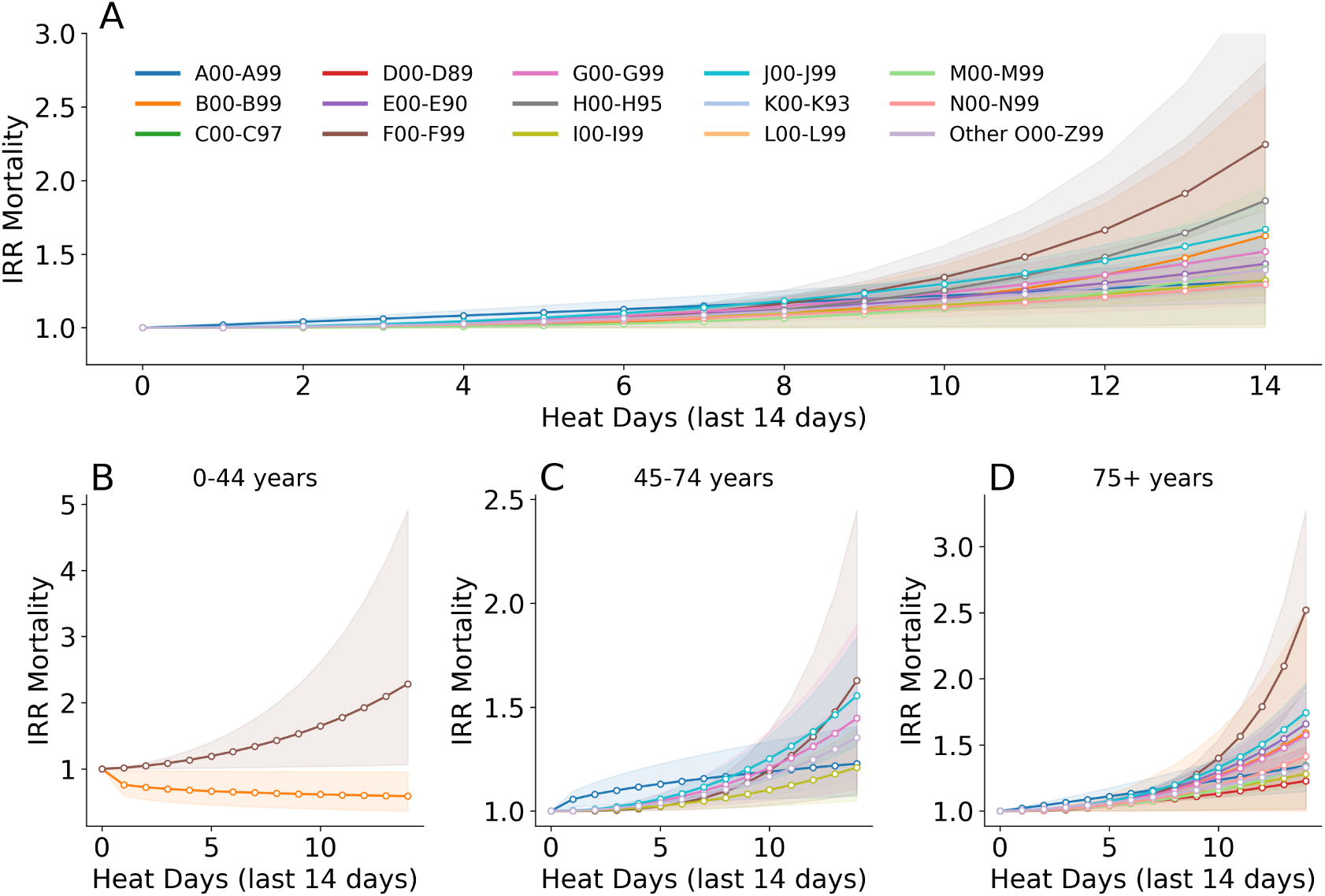
Impact of heat days (Tmax > 30°C) on in-hospital mortality, by diagnosis group and demographic strata. (A) IRRs for in-hospital mortality per diagnosis group (ICD-10 chapters A to N and O–Z combined) associated with the number of heat days (daily maximum temperature above 30 degrees) in the past 14 days. (B–D) IRRs for in-hospital mortality per diagnosis group stratified by age group: 0–44 years (B), 45–74 years (C), and 75+ years (D). Only statistically significant results are shown. Shaded bands represent 95% confidence intervals. Note the different scales in each panel.

**Table 8:**
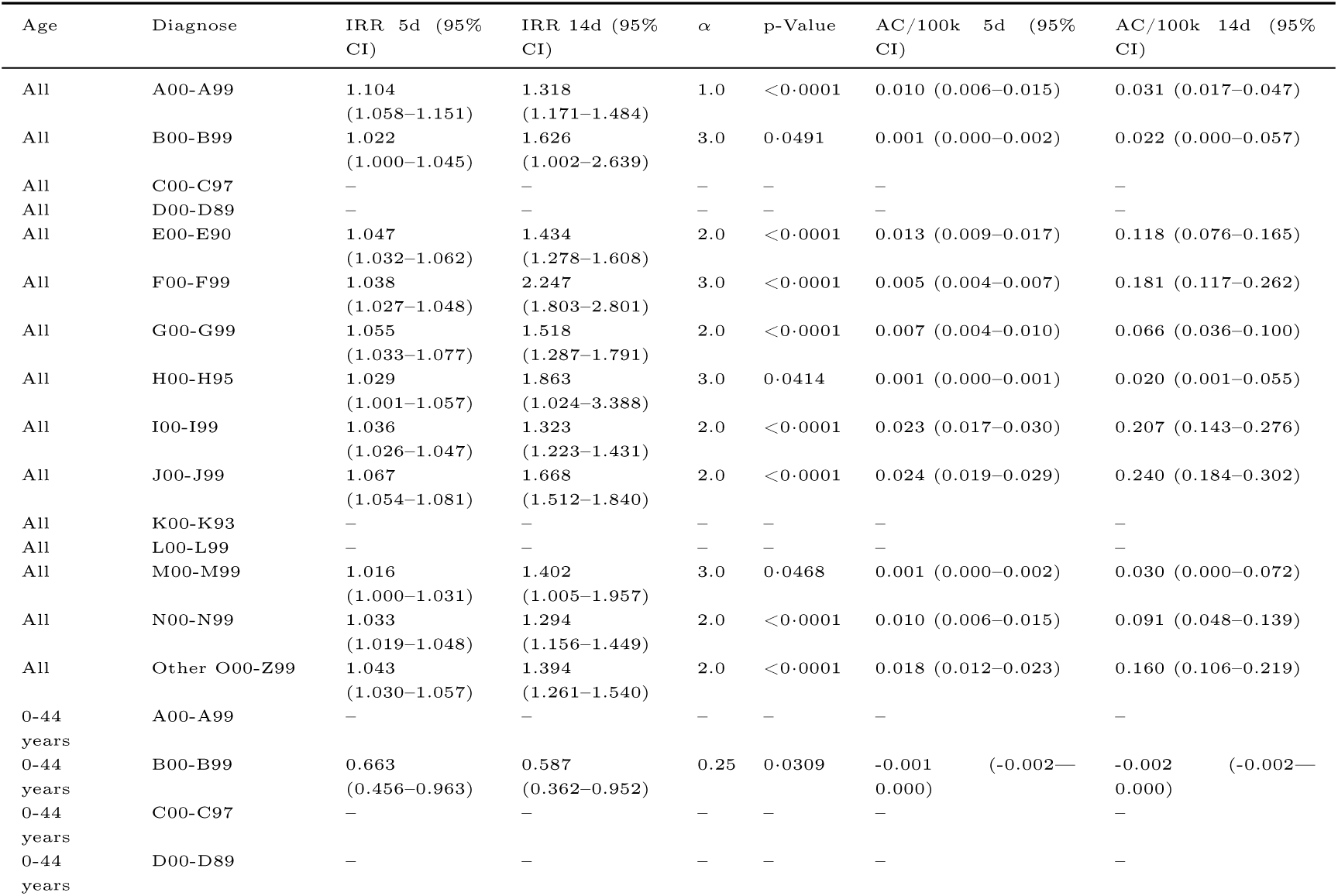
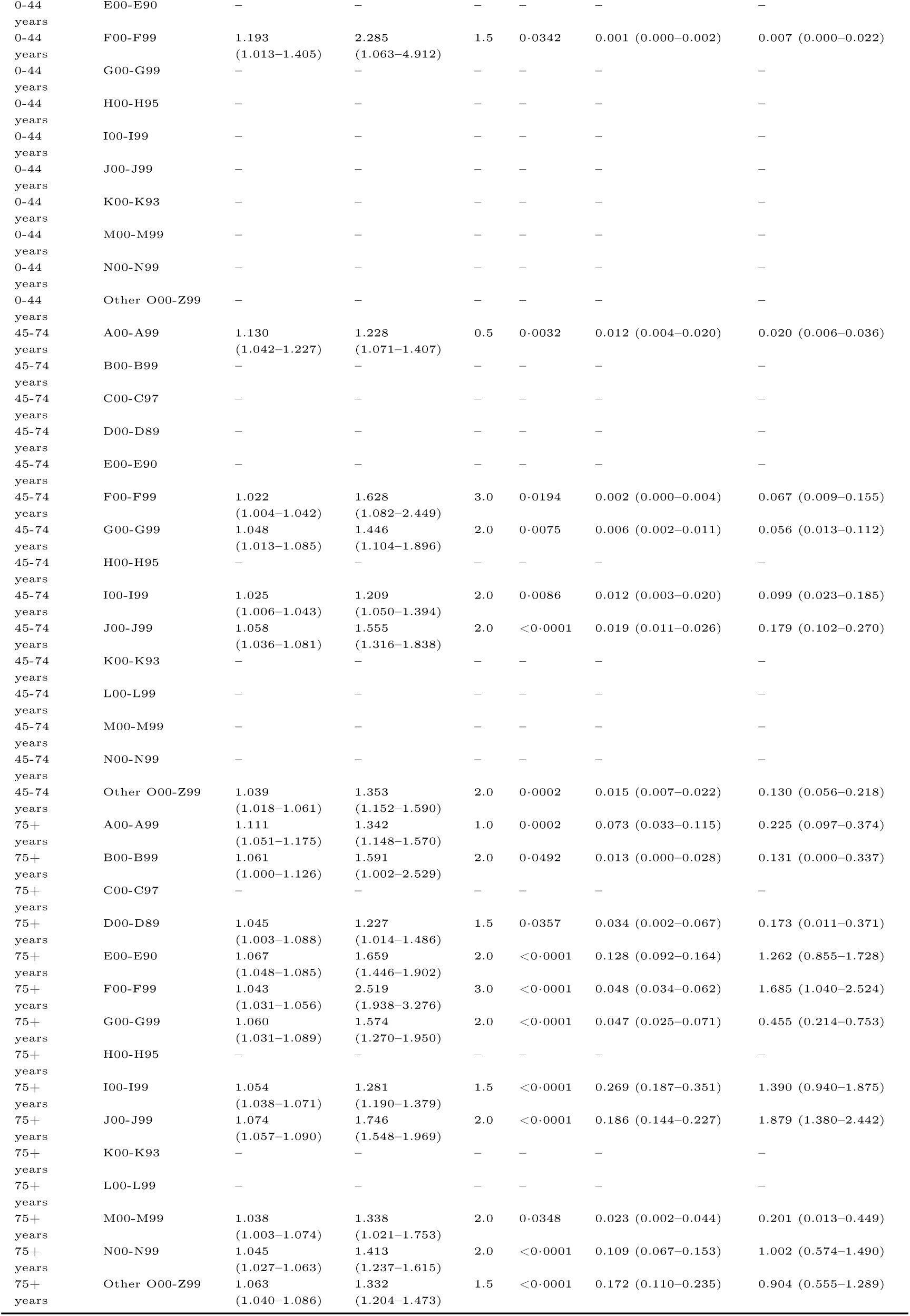
Estimated IRRs and additional cases (AC) per 100k with 95% CI by age group and diagnosis group at heat day definition Tmax > 30°C for outcome mortality (MT).

**Figure 17:**
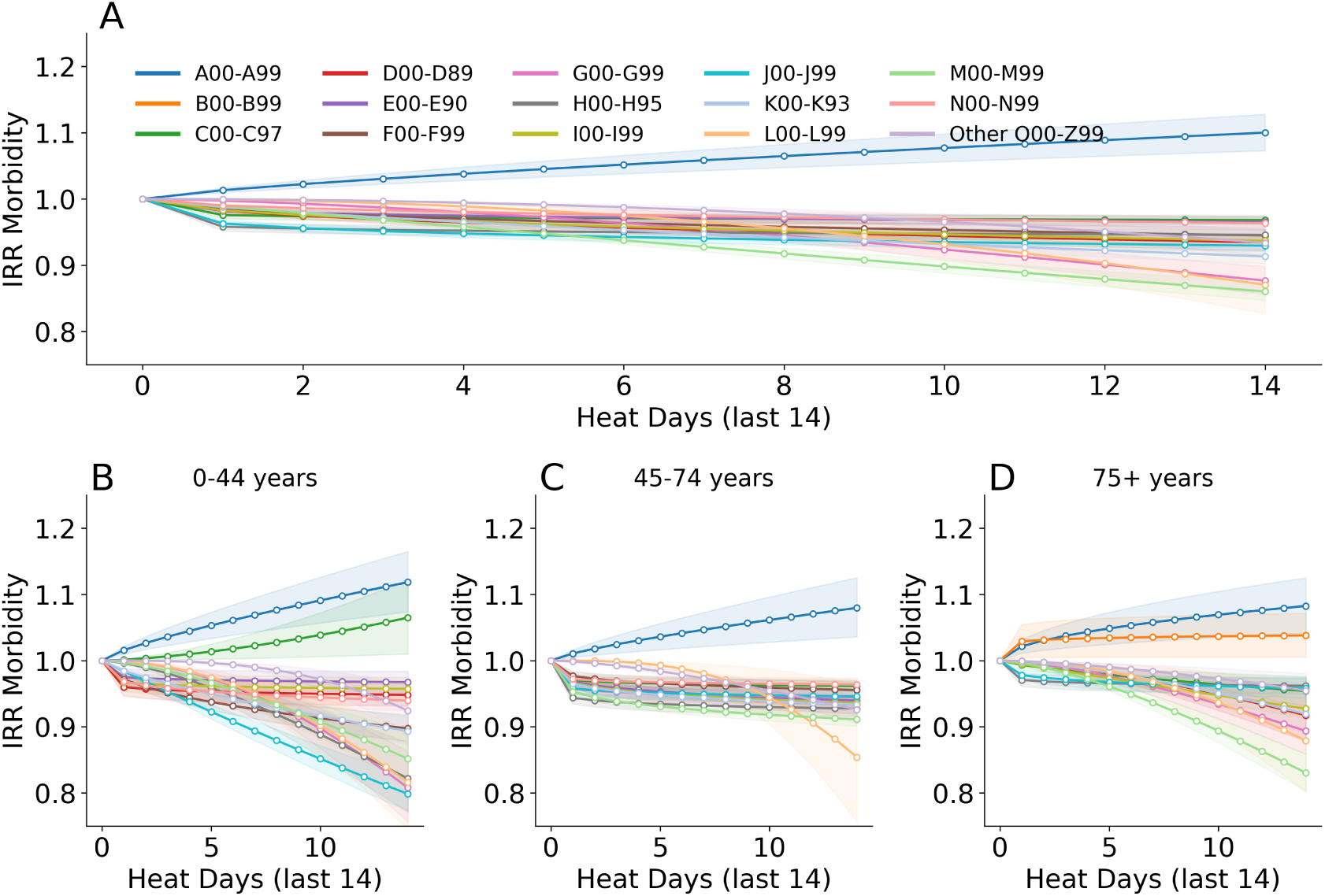
Impact of heat days (Tmax > 30°C) on hospital admissions, by diagnosis group and demographic strata.(A) IRRs for hospital admissions per diagnosis group (ICD-10 chapters A to N and O–Z combined) associated with the number of heat days (daily maximum temperature above 30 degrees) in the past 14 days. (B–D) IRRs for hospital admissions per diagnosis group associated with the number of heat days stratified by age group: 0–44 years (B), 45–74 years (C), and 75+ years (D). Only statistically significant results are shown. Shaded areas represent 95% confidence intervals.

**Table 9:**
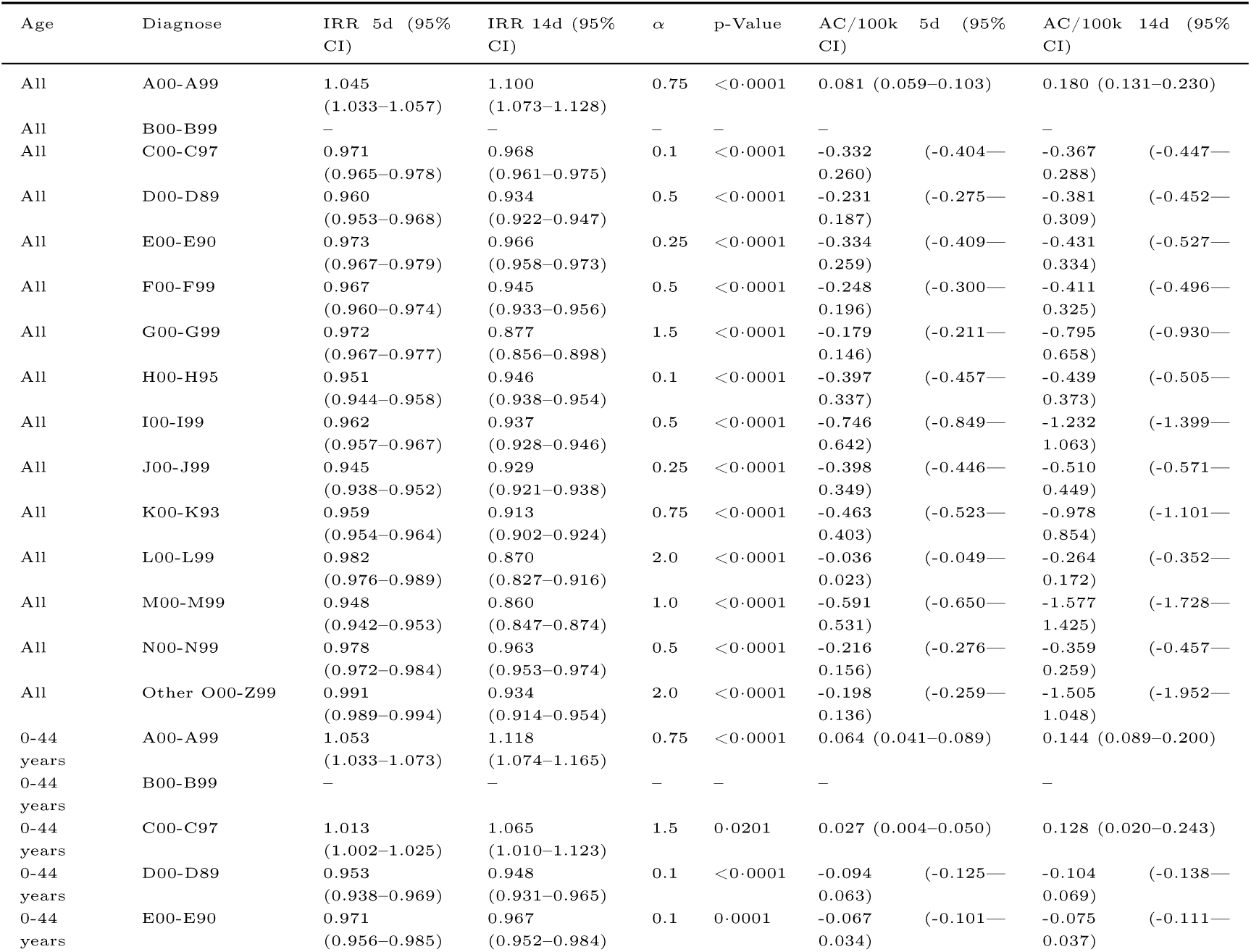
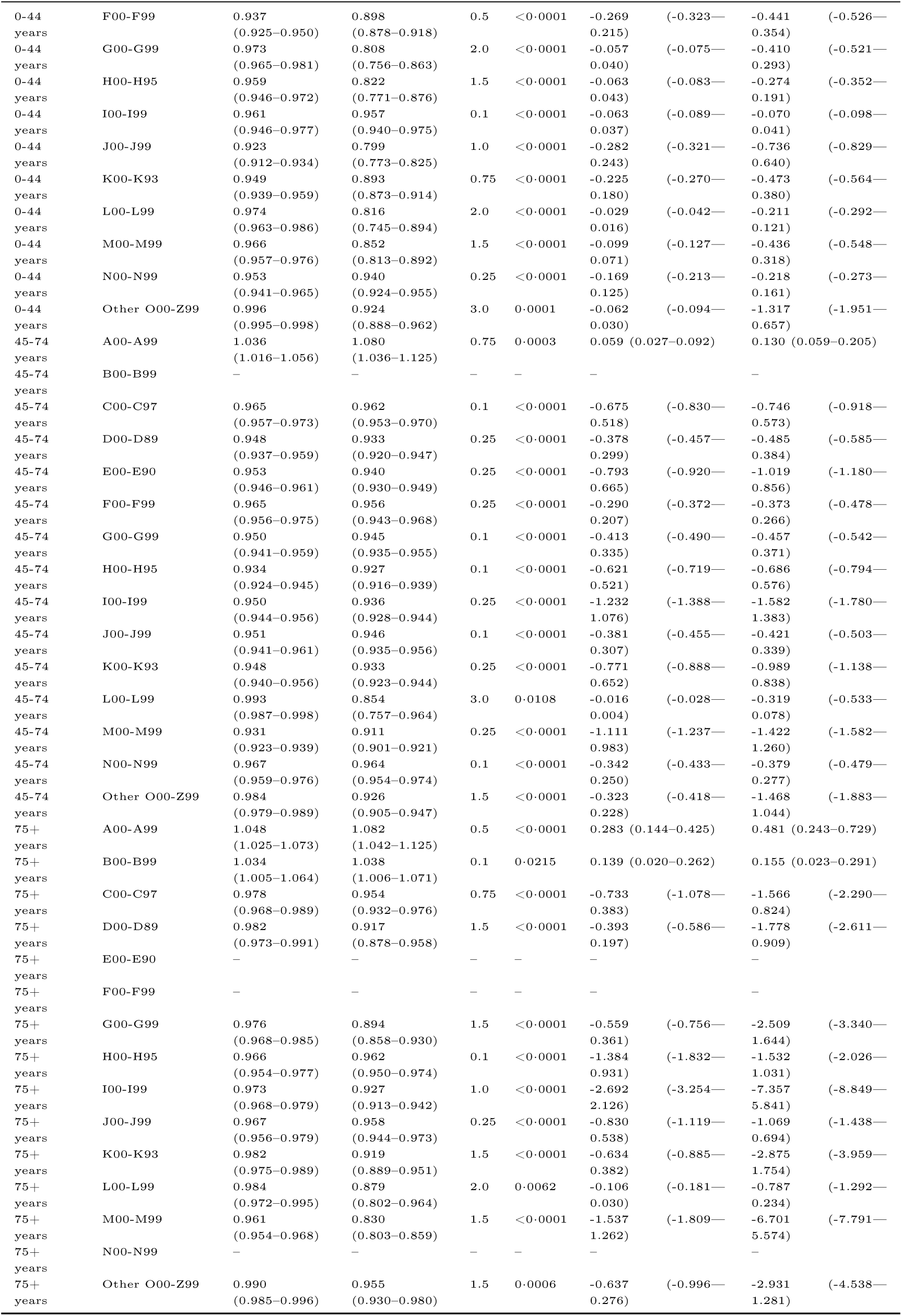
Estimated IRRs and additional cases (AC) per 100k with 95% CI by age group and diagnosis group at heat day definition Tmax > 30°C for outcome admissions (MB).

## References

[1] J. Hansen, M. Sato, R. Ruedy, K. Lo, D. W. Lea & M. Medina-Elizade, ‘Global temperature change,’ en, Proceedings of the National Academy of Sciences, 103, no. 39, pp. 14 288–14 293, Sep. 2006, issn: 0027-8424, 1091-6490. doi: 10.1073/pnas.0606291103. Accessed: 21st Aug. 2025. [Online]. Available: https://pnas.org/doi/full/10.1073/pnas.0606291103.

[2] K. Brugger, A. E. Schmidt & J. Delcour, ‘Factsheet: Krankenhausaufenthalte im direkten Zusam- menhang mit Hitze und Sonnenlicht in österreich (2002–2020),’ de, *Factsheet*, Gesundheit Österreich, 2022. [Online]. Available: https://jasmin.goeg.at/id/eprint/2279/1/Krankenhausaufenthalte%5C%20im%5C%20direkten%5C%20Zusammenhang%5C%20mit%5C%20Hitze%5C%20und%5C%20Sonnenlicht%5C%20in%5C%20%C3%96sterreich%5C_bf.pdf.

[3] D. Huppmann, M. Keiler, K. Riahi & H. Rieder, ‘Second Austrian Assessment Report on Climate Change | AAR2 - Full Report,’ en, Verlag der Österreichischen Akademie der Wissenschaften, Tech. Rep., Jun. 2025. doi: 10.1553/aar2. Accessed: 19th Aug. 2025. [Online]. Available: https://www.austriaca.at/?arp=0x00406e29.

[4] J. Ballester et al., ‘Heat-related mortality in Europe during the summer of 2022,’ en, Nature Medicine, 29, no. 7, pp. 1857–1866, Jul. 2023, issn: 1078-8956, 1546-170X. doi: 10.1038/s41591-023-02419-z. Accessed: 8th Jul. 2024. [Online]. Available: https://www.nature.com/articles/s41591-023-02419-z.

[5] K. L. Ebi et al., ‘Hot weather and heat extremes: Health risks,’ en, The Lancet, 398, no. 10301, pp. 698–708, Aug. 2021, issn: 0140-6736. doi: 10.1016/s0140-6736(21) 01208-3. Accessed: 9th Jul. 2025. [Online]. Available: https://linkinghub.elsevier.com/retrieve/pii/S0140673621012083.

[6] K. Brooks, O. Landeg, S. Kovats, M. Sewell & E. OConnell, ‘Heatwaves, hospitals and health system resilience in England: A qualitative assessment of frontline perspectives from the hot summer of 2019,’ en, BMJ Open, 13, no. 3, e068298, Mar. 2023, issn: 2044-6055, 2044-6055. doi: 10.1136/bmjopen-2022-068298. Accessed: 22nd Sep. 2025. [Online]. Available: https://bmjopen.bmj.com/lookup/doi/10.1136/bmjopen-2022-068298.

[7] H.-P. Hutter, H. Moshammer, P. Wallner, B. Leitner & M. Kundi, ‘Heatwaves in Vienna: Effects on mortality,’ en, Wiener klinische Wochenschrift, 119, no. 7-8, pp. 223–227, May 2007, issn: 0043- 5325, 1613-7671. doi: 10.1007/s00508-006-0742-7. Accessed: 19th Aug. 2025. [Online]. Available: http://link.springer.com/10.1007/s00508-006-0742-7.

[8] A. Matzarakis, S. Muthers & E. Koch, ‘Human biometeorological evaluation of heat-related mortality in Vienna,’ en, Theoretical and Applied Climatology, 105, no. 1-2, pp. 1–10, Aug. 2011, issn: 0177-798X, 1434-4483. doi: 10.1007/s00704-010-0372-x. Accessed: 8th Jul. 2024. [Online]. Available: http://link.springer.com/10.1007/s00704-010-0372-x.

[9] M. Hagen & P. Weihs, ‘Mortality during Heatwaves and Tropical Nights in Vienna between 1998 and 2022,’ en, Atmosphere, 14, no. 10, p. 1498, Sep. 2023, issn: 2073-4433. doi: 10.3390/atmos14101498. Accessed: 8th Jul. 2024. [Online]. Available: https://www.mdpi.com/2073-4433/14/10/1498.

[10] J. Liu et al., ‘Heat exposure and cardiovascular health outcomes: A systematic review and meta- analysis,’ 6, no. e484–95, 2022. doi: 10.1016/S2542-5196(22)00117-6.

[11] K. Knowlton, et al., ‘The 2006 California Heat Wave: Impacts on Hospitalizations and Emergency Department Visits,’ en, Environmental Health Perspectives, 117, no. 1, pp. 61–67, Jan. 2009, issn: 0091-6765, 1552-9924. doi: 10 .1289/ehp .11594. Accessed: 24th Apr. 2024. [Online]. Available: https://ehp.niehs.nih.gov/doi/10.1289/ehp.11594.

[12] D. Oudin Åström, F. Bertil & R. Joacim, ‘Heat wave impact on morbidity and mortality in the elderly population: A review of recent studies,’ en, Maturitas, 69, no. 2, pp. 99–105, Jun. 2011, issn: 03785122. doi: 10.1016/j.maturitas.2011.03.008. Accessed: 24th Apr. 2024. [Online]. Available: https://linkinghub.elsevier.com/retrieve/pii/S0378512211000806.

[13] R. S. Kovats, S. Hajat & P. Wilkinson, ‘Contrasting patterns of mortality and hospital admissions during hot weather and heat waves in Greater London, UK,’ en, Occupational and Environmental Medicine, 61, no. 11, pp. 893–898, Nov. 2004, issn: 1351-0711, 1470-7926. doi: 10.1136/oem.2003.012047. Accessed: 22nd Aug. 2025. [Online]. Available: https://oem.bmj.com/lookup/doi/10.1136/oem.2003.012047.

[14] G. A. Robertson, A. G. Marsh, S. L. Gill, D. Martin, D. J. Lowe & B. Jamal, ‘The influence of heatwave temperatures on fracture patient presentation to hospital,’ en, Injury, 53, no. 10, pp. 3163– 3171, Oct. 2022, issn: 00201383. doi: 10.1016/j.injury.2022.07.007. Accessed: 22nd Aug. 2025. [Online]. Available: https://linkinghub.elsevier.com/retrieve/pii/S0020138322004703.

[15] M. Nitschke, G. R. Tucker & P. Bi, ‘Morbidity and mortality during heatwaves in metropolitan Adelaide,’ en, Medical Journal of Australia, 187, no. 11-12, pp. 662–665, Dec. 2007, issn: 0025-729X, 1326-5377. doi: 10.5694/j.1326-5377.2007.tb01466.x. Accessed: 22nd Aug. 2025. [Online]. Available: https://onlinelibrary.wiley.com/doi/10.5694/j.1326-5377.2007.tb01466.x.

[16] C. J. Gronlund, A. Zanobetti, J. D. Schwartz, G. A. Wellenius & M. S. O’Neill, ‘Heat, Heat Waves, and Hospital Admissions among the Elderly in the United States, 1992–2006,’ en, Environmental Health Perspectives, 122, no. 11, pp. 1187–1192, Nov. 2014, issn: 0091-6765, 1552-9924. doi: 10. 1289/ehp.1206132. Accessed: 22nd Aug. 2025. [Online]. Available: https://ehp.niehs.nih.gov/doi/10.1289/ehp.1206132.

[17] H. Achebak et al., ‘Heat Exposure and Cause-Specific Hospital Admissions in Spain: A Nationwide Cross-Sectional Study,’ en, Environmental Health Perspectives, 132, no. 5, p. 057 009, May 2024, issn: 0091-6765, 1552-9924. doi: 10.1289/EHP13254. Accessed: 22nd Apr. 2026. [Online]. Available: https://ehp.niehs.nih.gov/doi/10.1289/EHP13254.

[18] A. M. Alho, A. P. Oliveira, S. Viegas & P. Nogueira, ‘Effect of heatwaves on daily hospital admissions in Portugal, 2000–18: An observational study,’ en, The Lancet Planetary Health, 8, no. 5, e318– e326, May 2024, issn: 2542-5196. doi: 10.1016/s2542-5196(24)00046-9. Accessed: 14th Jul. 2025. [Online]. Available: https://linkinghub.elsevier.com/retrieve/pii/S2542519624000469.

[19] M. Karlsson & N. R. Ziebarth, ‘Population health effects and health-related costs of extreme temperatures: Comprehensive evidence from Germany,’ en, Journal of Environmental Economics and Management, 91, pp. 93–117, Sep. 2018, issn: 00950696. doi: 10.1016/j.jeem.2018.06.004. Accessed: 21st Apr. 2026. [Online]. Available: https://linkinghub.elsevier.com/retrieve/pii/S0095069616304636.

[20] P. Fahr et al., ‘Quantifying the health-care burden of temperature in the National Health Service in England: An economic analysis of resource use and costs,’ en, The Lancet Planetary Health, 9, no. 12, p. 101 373, Dec. 2025, issn: 25425196. doi: 10.1016/j.lanplh.2025.101373. Accessed: 22nd Apr. 2026. [Online]. Available: https://linkinghub.elsevier.com/retrieve/pii/S2542519625002517.

[21] C. F. Gould et al., ‘Temperature extremes impact mortality and morbidity differently,’ en, Science AdvAnceS, 2025.

[22] F. Rubel, K. Brugger, K. Haslinger & I. Auer, ‘The climate of the European Alps: Shift of very high resolution Köppen-Geiger climate zones 1800–2100,’ en, Meteorologische Zeitschrift, 26, no. 2, pp. 115–125, Apr. 2017, issn: 0941-2948. doi: 10.1127/metz/2016/0816. Accessed: 19th Aug. 2025. [Online]. Available: http://www.schweizerbart.de/papers/metz/detail/26/87237/The_climate_of_the_European_Alps_Shift_of_very_hig?af=crossref.

23. A. Gasparrini, ‘Distributed lag linear and non-linear models for time series data,’ en, 2013.

[24] H. Achebak, D. Devolder & J. Ballester, ‘Heat-related mortality trends under recent climate warming in Spain: A 36-year observational study,’ en, PLOS Medicine, 15, no. 7, J. A. Patz, Ed., e1002617, Jul. 2018, issn: 1549-1676. doi: 10.1371/journal.pmed.1002617. Accessed: 22nd Apr. 2026. [Online]. Available: https://dx.plos.org/10.1371/journal.pmed.1002617.

[25] E. Gallo et al., ‘Heat-related mortality in Europe during 2023 and the role of adaptation in protecting health,’ Nature Medicine, 30, no. 11, pp. 3101–3105, Nov. 2024, issn: 1546-170X. doi: 10.1038/s41591-024-03186-1. [Online]. Available: 10.1038/s41591-024-03186-1.

[26] X. Basagaña et al., ‘Heat Waves and Cause-specific Mortality at all Ages,’ en, Epidemiology, 22, no. 6, pp. 765–772, Nov. 2011, issn: 1044-3983. doi: 10.1097/ede.0b013e31823031c5. Accessed: 10th Jul. 2025. [Online]. Available: https://journals.lww.com/00001648-201111000-00002.

[27] S. Hajat & T. Kosatky, ‘Heat-related mortality: A review and exploration of heterogeneity,’ en, Journal of Epidemiology & Community Health, 64, no. 9, pp. 753–760, Sep. 2010, issn: 0143-005X. doi: 10.1136/jech.2009.087999. Accessed: 22nd Aug. 2025. [Online]. Available: https://jech.bmj.com/lookup/doi/10.1136/jech.2009.087999.

[28] G. Rey et al., ‘The impact of major heat waves on all-cause and cause-specific mortality in France from 1971 to 2003,’ en, International Archives of Occupational and Environmental Health, 80, no. 7, pp. 615–626, Jul. 2007, issn: 0340-0131, 1432-1246. doi: 10.1007/s00420-007-0173-4. Accessed: 22nd Aug. 2025. [Online]. Available: https://link.springer.com/10.1007/s00420-007-0173-4.

[29] B. G. Anderson & M. L. Bell, ‘Weather-Related Mortality: How Heat, Cold, and Heat Waves Affect Mortality in the United States,’ en, Epidemiology, 20, no. 2, pp. 205–213, Mar. 2009, issn: 1044-3983. doi: 10.1097/EDE.0b013e318190ee08. Accessed: 22nd Aug. 2025. [Online]. Available: https://journals.lww.com/00001648-200903000-00011.

[30] B. Hoffmann, S. Hertel, T. Boes, D. Weiland & K.-H. Jöckel, ‘Increased Cause-Specific Mortality Associated with 2003 Heat Wave in Essen, Germany,’ en, Journal of Toxicology and Environmental Health, Part A, 71, no. 11-12, pp. 759–765, Jun. 2008, issn: 1528-7394, 1087-2620. doi: 10.1080/15287390801985539. Accessed: 22nd Aug. 2025. [Online]. Available: http://www.tandfonline.com/doi/abs/10.1080/15287390801985539.

[31] R. Kaiser, A. Le Tertre, J. Schwartz, C. A. Gotway, W. R. Daley & C. H. Rubin, ‘The Effect of the 1995 Heat Wave in Chicago on All-Cause and Cause-Specific Mortality,’ en, American Journal of Public Health, 97, no. Supplement_1, S158–S162, Apr. 2007, issn: 0090-0036, 1541-0048. doi: 10.2105/AJPH.2006.100081. Accessed: 22nd Aug. 2025. [Online]. Available: https://ajph.aphapublications.org/doi/full/10.2105/AJPH.2006.100081.

[32] G. Mastrangelo, U. Fedeli, C. Visentin, G. Milan, E. Fadda & P. Spolaore, ‘Pattern and determinants of hospitalization during heat waves: An ecologic study,’ en, BMC Public Health, 7, no. 1, p. 200, Dec. 2007, issn: 1471-2458. doi: 10.1186/1471-2458-7-200. Accessed: 22nd Aug. 2025. [Online]. Available: https://bmcpublichealth.biomedcentral.com/articles/10.1186/1471-2458-7-200.

[33] M. Stafoggia et al., ‘Factors affecting in-hospital heat-related mortality: A multi-city case-crossover analysis,’ en, Journal of Epidemiology & Community Health, 62, no. 3, pp. 209–215, Mar. 2008, issn: 0143-005X. doi: 10.1136/jech.2007.060715. Accessed: 22nd Aug. 2025. [Online]. Available: https://jech.bmj.com/lookup/doi/10.1136/jech.2007.060715.

[34] R. V. Remigio et al., ‘Association of Extreme Heat Events With Hospital Admission or Mortality Among Patients With End-Stage Renal Disease,’ en, JAMA Network Open, 2, no. 8, e198904, Aug. 2019, issn: 2574-3805. doi: 10.1001/jamanetworkopen.2019.8904. Accessed: 22nd Aug. 2025. [Online]. Available: https://jamanetwork.com/journals/jamanetworkopen/fullarticle/2747697.

